# Long-term PM_2.5_ exposure and depressive symptoms in China: a quasi-experimental study

**DOI:** 10.1101/2020.07.07.20147959

**Authors:** Tao Xue, Tianjia Guan, Yixuan Zheng, Guannan Geng, Qiang Zhang, Yao Yao, Tong Zhu

## Abstract

**Background:** Air pollutants, particularly fine particulate matters (**PM**_2.5_) have been associated with mental disorder such as depression. Clean air policy (CAP, *i*.*e*., a series of emission-control actions) has been shown to reduce the public health burden of air pollutions. There were few studies on the health effects of CAP on mental health, particularly, in low-income and middle-income countries (LMICs). We investigated the association between a stringent CAP and depressive symptoms among general adults in China.

**Methods:** We used three waves (2011, 2013 and 2015) of the China Health and Retirement Longitudinal Study (CHARLS), a prospective nationwide cohort of the middle-aged and older population in China. We assessed exposure to **PM**_2.5_ through a satellite-retrieved dataset. We implemented a difference-in-differences (**DID**) approach, under the quasi-experimental framework of the temporal contrast between 2011 (before the CAP) and 2015 (after the CAP), to evaluate the effect of CAP on depressive symptoms. The association was further explored using a mixed-effects model of the three waves. To increase the interpretability, the estimated impact of **PM**_2.5_ was compared to that of aging, an established risk factor for depression.

**Findings:** Our analysis included 15,954 participants. In the **DID** model, we found a 10-μg/m^3^ reduction of **PM**_2.5_ concentration was associated with a 4.14% (95% CI: 0.41–8.00%) decrement in the depressive score. The estimate was similar to that from the mixed-effects model (3.63% [95% CI, 2.00–5.27%]). We also found improved air quality during 2011-2015 offset the negative impact from 5-years’ aging.

**Interpretation:** The findings suggest that implementing CAP may improve mental wellbeing of adults in China and other LMICs.

**Funding:** National Natural Science Foundation and Ministry of Science and Technology of China, and Energy Foundation.

## Introduction

Mental disorders have contributed to a large proportion of the global burden of diseases by causing either non-fatal (e.g., disability) or fatal outcomes (e.g., suicide). Mental illness is the leading cause of years lived with disability (YLD), and the related disease burden increased by 37.6% from 1990 to 2010.^1^ The increased burden of mental illness may be caused by both demographic dynamics, and changes in relevant risk factors, such as inactive lifestyles and environmental pollutants. For instance, 37.5% of the increase in depression-associated YLDs during 1990-2010 were attributable to population growth and ageing^2^. Recent epidemiological evidence suggests that air pollution may adversely affect mental health,^3-10^ and thus increase the disability burden.^11^ Due to the ubiquitous and prolonged exposure to ambient pollutants, particularly in low-income and middle-income countries (LMICs), air quality can be a major contributor to mental illness, such as depression.^12^ In addition, some LMICs, such as China, started to implement a series of emission-control policies, which are usually known as the clean air policy (CAP). However, few studies examined the effects of CAP on mental health, particularly in LMICs.

To address severe air pollution issues and protect public health, the State Council of China promulgated the toughest-ever air pollution control on September 10, 2013,^13-16^ including optimization of the industrial structure, improvements in end-of-pipe control, and reductions in residential usage of unclean fuels. The policy is officially named as Air Pollution Prevention and Control Action Plan. It is comparable with similar policies in developed countries (*e*.*g*., United States), and is thus referred as CAP for short, hereafter. The long-term concentrations of fine particulate matter less than 2.5 µm in diameter (**PM**_2.5_), the major air pollutant in China, decreased rapidly nationwide from 67.4 μg/m^3^ in 2013 to 45.5 μg/m^3^ in 2017.^15^ This dramatic change in air quality provides an opportunity to study the mental effect of **PM**_2.5_ under a quasi-experimental scenario. A quasi-experiment can result in a shape contrast of air pollution levels during a relatively short period, which is advantageous to indicate causal by controlling unmeasured and omitted confounders.^17,18^ In this study, we evaluated the association between long-term **PM**_2.5_ exposure and depressive symptoms, under the quasi-experimental framework of the temporal contrast between 2011 (before the actions) and 2015 (after the actions). We made use of the China Health and Retirement Longitudinal Study (CHARLS), which repeatedly measured the depressive score (CES-D-10, 10-item of Center for Epidemiologic Studies Depression Scale) of a representative sample of middle-aged and older Chinese adults in 2011, 2013, and 2015.^19^ Due to the overlap between the CHARLS study period and the duration of China’s clean air actions, we were able to associate the reduction of concentrations of **PM**_2.5_ to depressive score changes at the individual level, and to quantify the impact of improved air quality on adult metal health. Because the CES-D-10 is not clinically indicative, interpreting the impact of air quality is difficult. To increase interpretability, we further compared the effect of **PM**_2.5_ concentrations to that of human aging, an established risk factor for poor mental health.^1,2,20^

## Methods

### Study population

The studied adults were obtained from the China Health and Retirement Longitudinal Study (h CHARLS).^19,21^ CHARLS is an ongoing nationwide longitudinal survey on the health and socioeconomic status of middle-aged and older Chinese adults. Using a four-stage and well-established sample design, CHARLS researchers recruited a representative sample of ∼20,000 Chinese adults from ∼150 county-level units in 2011 (Figure 1). Subsequently, well-trained fieldworkers regularly visited the respondents every 2 years (Figure S1), and thus surveyed a national-scale cohort of Chinese adults. In each survey wave, all subjects were visited by well-trained interviewers in a face-to-face computer-assisted personal interview, which gathered data on demographic characteristics, behavioral risk factors, the residential city, and housing conditions. Details of the study design and the purpose of the CHALRS are provided elsewhere.^19^ All procedures involving human subjects/patients were approved by the Ethics Review Committee of Peking University (IRB00001052–11015). CHARLS has supported studies on mental health risk factors^22^ and the effects of air pollution on other outcomes.^23^ This study utilized the open-accessed CHARLS data, which is publicly available from the website: http://opendata.pku.edu.cn/. All analyses in this study adhered to the data usage guidelines.

**Figure 1.**
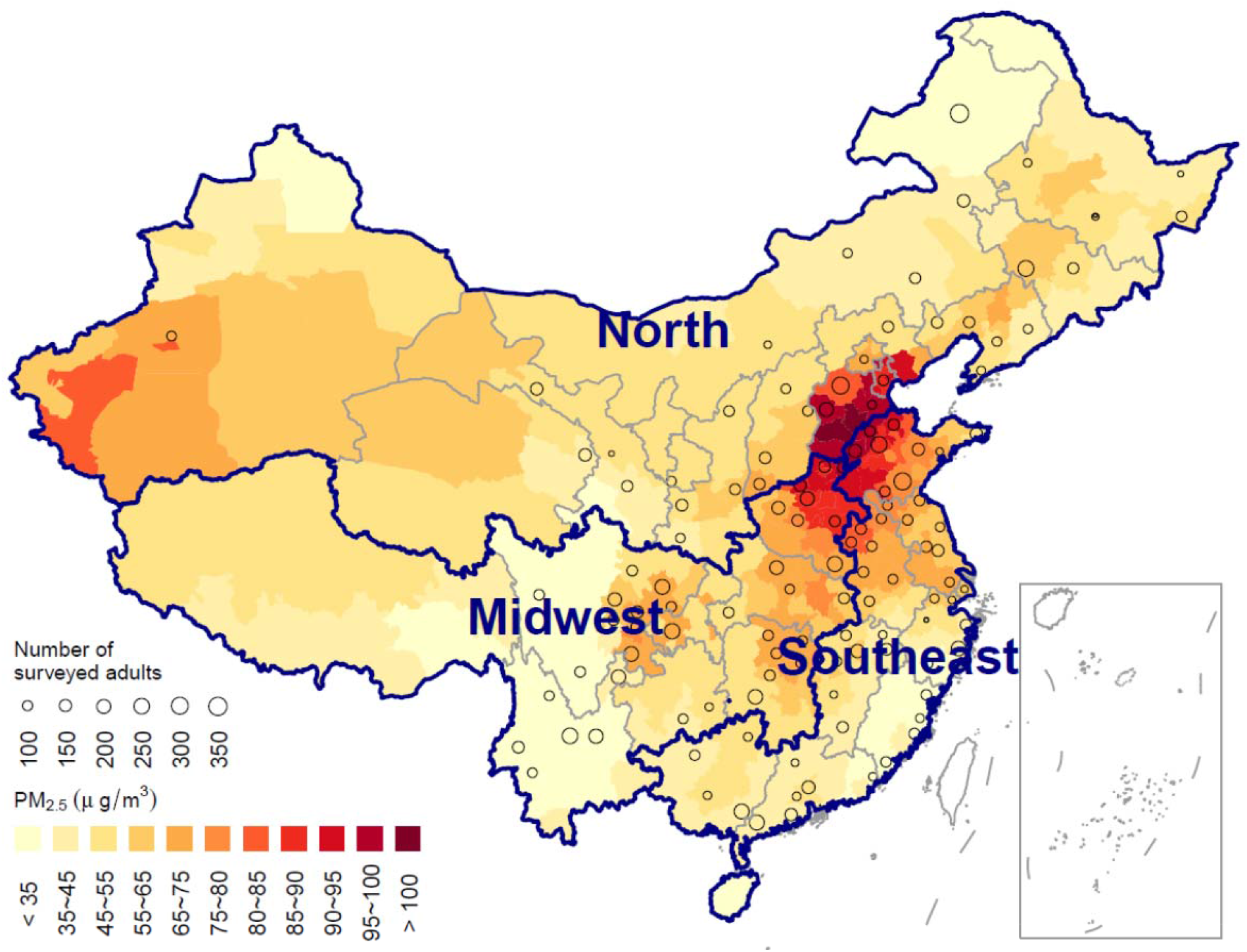
Map of study region with long-term averages of PM_2.5_ concentrations (2010–2015).

This study was based on currently available data from CHARLS surveys in 2011, 2013, and 2015, which involved a sample of 24,805 individual adults. To conduct a longitudinal analysis, we focused on the valid records of older adults (≥ 40 years old), who were visited at least twice. A total of 16,151 adults remained after excluding subjects who were only observed once. We further excluded the adults without age data (which was derived from the time of the survey and the birthdate) and adults <40 years old at survey time. Finally, this study involved 41,031 observations of 15,954 individual adults distributed across 447 communities in 126 cities and counties (Figure 1). The data cleaning diagram is displayed in Figure S1.

### Environmental exposure

The ambient exposure assessment was based on the **PM**_2.5_ Hindcast Database for China (2000–2016) introduced in our previous work.^24^ The database can be accessed from the website: http://www.meicmodel.org/dataset-phd.html. Due to the lack of nationwide monitoring data on **PM**_2.5_ concentrations in China before 2013, historical **PM**_2.5_ concentrations were estimated from satellite remote-sensing measurements and outputs from a chemical transport model (CTM) of air pollution emission inventories. Satellite sensors, such as the Moderate Resolution Imaging Spectroradiometer, retrieve the column concentrations of aerosols from the earth’s surface to the top of the atmosphere by measuring electromagnetic signals. Satellite-based annual estimates of **PM**_2.5_ have been applied in many health-related studies at the national and global scales.^13,25^ The CTM simulations were based on the multi-resolution emission inventory for China (http://www.meicmodel.org/index.html). The results provide a complete characterization of the spatiotemporal variations in **PM**_2.5_ concentrations and have been applied to support studies on health risk assessments and relevant policy analyses in China.^16,26^ In a previous study,^24^ we developed a machine-learning model to bring the satellite measurements and CTM simulations together by relating them to the nationwide monitoring concentrations of **PM**_2.5_ from 2013 to 2016, and then applied the model to hindcast the **PM**_2.5_ values before 2013. The estimator was in good agreement with the *in-situ* observations on monthly (R^2^ = 0.71) and yearly scales (R^2^ = 0.77) and has been utilized in other epidemiological studies. Please refer to our previous study for more details on the **PM**_2.5_ estimator.^24^

The original **PM**_2.5_ data have a spatial resolution of 0.1° × 0.1° and daily concentrations across the mainland of China, during 2000–2016. For consistency, exposure assessments before and after 2013 were based on the estimated **PM**_2.5_ concentrations. The subjects in the CHARLS could only be geo-coded to each participant’s regionalization code due to confidential reason. Therefore, we first pooled the **PM**_2.5_ data into city-level averages by matching the pixels of a regular grid with a map of China’s prefectures (Figure 1), and further calculated the monthly averages. We utilized the **PM**_2.5_ concentration averages during the 12 months preceding the surveyed months as the exposure values (Figure S1).

We also obtained gridded estimates of temperature with an original resolution of 0.1° × 0.1° by fusing the satellite measurements of land surface temperatures, *in-situ* observations, and simulations from a weather-forecast research model.^3^ The three types of temperature values were assembled by day using a universal kriging approach. Random cross-validations indicated that the fused estimates were in good agreement with the monitored values (R^2^ = 0.96). Details of the temperature data assembly are documented in our previous study.^3^ City-level monthly averages for temperature data were also calculated for each record before the regression analyses.

### Measurement of depressive symptoms

The mental health status of the subjects was measured using the 10-item Center for Epidemiologic Studies Depression scale (CES-D-10). The validity of the CES-D-10 has been examined among Chinese adults.^27^ Each question measures the frequency of a specific type of negative mood (e.g., fearful or depressed) using a score of 0 (rarely or none), 1 (some days), 2 (occasionally), or 3 (most of the time). The CES-D-10 questions were included into the standard CHARLS questionnaires, which were collected during a computer-assisted interview by fieldworkers. We calculated the sum of all question-specific scores as the depressive score (CES-D-10) to indicate the general status of depressive symptoms for each subject. The depressive score ranges from 0 to 30, with a higher score indicating a higher severity of depressive symptoms.

### Statistical analyses

We applied multiple methods to evaluate the effect of CAP and to derive the association between **PM**_2.5_ and CES-D-10. Relationships of the methods and their different properties are presented in Figure S1.

First, we utilized the difference-in-difference (DID) approach, the typical model in quasi-experiments. The conventional DID model was designed for binary exposure (as illustrated in the preliminary analysis, Figure S2). Because **PM**_2.5_ is a continuous variable, a regression analysis of the association between changes in **PM**_2.5_ and CES-D-10 score was performed in our DID analysis. More details on the DID model are documented in Supplemental text (S1).

Next, to make full use of the data, we conducted a longitudinal analysis of the repeated measurements on CES-D-10 score and **PM**_2.5_ exposure using a mixed-effects model. Therefore, the modelling results were referred as longitudinal associations hereafter. The aim of the longitudinal analysis was to clarify the impact of **PM**_2.5_ exposure by comparing it with the impact of other risk factors.

Additionally, the fixed-effects model is a generalized version of the DID method that can incorporate more than two repeated measurements.^28^ It is also identical to the random-effects model, except for modelling subject-specific effects with a batch of dummy variables instead of random terms (Figure S1). A longitudinal regression with fixed effects is considered less biased, but also less efficient, than a comparable regression with random effects.^29^ The fixed-effects model was utilized to examine the robustness of the longitudinal associations.

Finally, the hypothesis test on whether CAP affected depression was relied on the DID model, because it directly mimics the scenarios with and without CAP (Figure S2), and thus is the canonical method to evaluate the effect of the policy. For the quantitative association between **PM**_2.5_ and CES-D-10 score, we relied on the mixed-effects model due to its efficacity and reliability. The overall conclusion was relied on the consistency between estimates from different models, by examining whether (1) the direction of association or its significance level was changed between models and (2) the estimated confidential intervals were overlapped with each other.

### Longitudinal association analyses

The mixed-effects model was specified as follows:

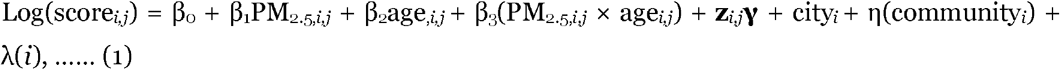

where *i* denotes the subject index; *j* denotes the visit index; β_0_ denotes the intercept; β_1-3_ denote the regression coefficients for **PM**_2.5_, age, and their interaction term, respectively; **z**_*i,j*_ denotes a set of adjusted covariates and **γ** denotes the corresponding coefficients; city_*i*_ denotes a fixed effect to control the unmeasured city-specific risk factors of depression, such as traditional culture ^30^; and λ and η denote two random slopes to model the correlations between records from the same subject or the same community, respectively. The adjusted covariates (**z**_*i,j*_) included (1) annual temperature; (2) demographic characteristics (urban/rural residency, sex, education level, and marriage status); (3) lifestyle risk factors (smoking and drinking); (4) housing conditions (cooking energy type, building type, residential rent payment, presence of an in-house telephone, and indoor temperature maintenance). The regression incorporating all the above covariates was termed the fully adjusted model. The model might be too complex to produce a stable estimator, and a large fraction of the regressed samples involved the imputed missing values, which increased the uncertainty of the model. To examine whether the estimated associations were sensitive to these limitations, besides the full adjustment, we also applied a standard adjustment, which involved only the first three sets of covariates, demographic characteristics, and lifestyle risk factors. The association was also evaluated by ER, similar to the DID model.

### Sensitivity analyses

In sensitivity analyses of the longitudinal model, we explored how the estimated association between **PM**_2.5_ concentration and the CES-D-10 score varied with (1) sub-regions of the study domain (Figure 1), (2) demographic sub-groups, and (3) exposure levels. Considering the balanced sample sizes (Figure 1), we divided the study domain into three sub-regions: Midwest (n = 13,886), North (n = 11,488) and Southeast (n = 15,657) China. We utilized interaction analyses to explore the modifications on the effect of **PM**_2.5_ concentrations by a sub-region indicator or demographic characteristic (e.g., sex), and replaced the linear term of **PM**_2.5_ concentration in the regression model with a penalized spline term to examine the linearity of the effect. We also conducted parallel analyses for the association between age and the CES-D-10 score to compare the effect of **PM**_2.5_ concentration to that of population aging. Finally, we also utilized a bootstrap method to evaluate how the lack of geographic addresses impacted the estimated association. To protect confidentiality, CHARLS only released the city-level geographic information, which could introduce exposure measurement errors into the association estimation. We applied a bootstrap method to incorporate uncertainty embedded in the data-generation procedure into the model estimation. Details of the bootstrap method were documented in supplemental text (S2).

### Assessment of impact of PM_2.5_

To illustrate the impact of the changes in air quality on depression, we conducted a post-hoc analysis based on the 9,123 adults who participated in all three CHARLS waves. We first calculated the change in one risk factor (Δ*x*_*i*_ = *x*_*i, 2011*_ − *x*_*i, 2015*_; *x* = **PM**_2.5_, age, or *z*) from 2011 to 2015, and quantified its impact on CES-D-10 score as [exp(Δ*x*_*i*_*β*_*x*_) − 1] × 100%. We compared the impact of the risk factors in the association model with standard adjustment, and focused on the combined impact of **PM**_2.5_ reduction and population aging in a group of adults.

All statistical analyses were performed using R (version 3.3.2; R Foundation for Statistical Computing, Vienna, Austria). The linear mixed-effects and fixed-effects models were inferred using the *lme4* package ^31^ and the *plm* package ^32^, respectively. Imputation was performed using the *mice* package ^33^. Inverse probability weights were calculated using the *ipw* package. Penalized spline functions were parameterized using the *mgcv* package ^34^, and inference of the nonlinear mixed-effects models was done using the *gamm4* package ^35^. The relevant R codes are documented in the Supplementary materials.

## Results

### Descriptive summary

This study involved 15,954 adults, and each was visited an average of 2.6 times. In 2011, the mean age of the adults was 58.4 years (standard deviation: 9.4 years). In the 2011, 2013, or 2015 CHARLS wave, the mean CES-D-10 score was 8.3, 7.8 or 8.1, and the corresponding concentration of **PM**_2.5_ was 61.6, 60.3 or 53.1 μg/m^3^, respectively. A summary of the demographic information of the adults in our study is presented in Table 1. The longitudinal variables, including environmental exposures, are summarized in Table 2. More detailed descriptions on the studied adults are documented in supplemental text (S3).

**Table 1.**
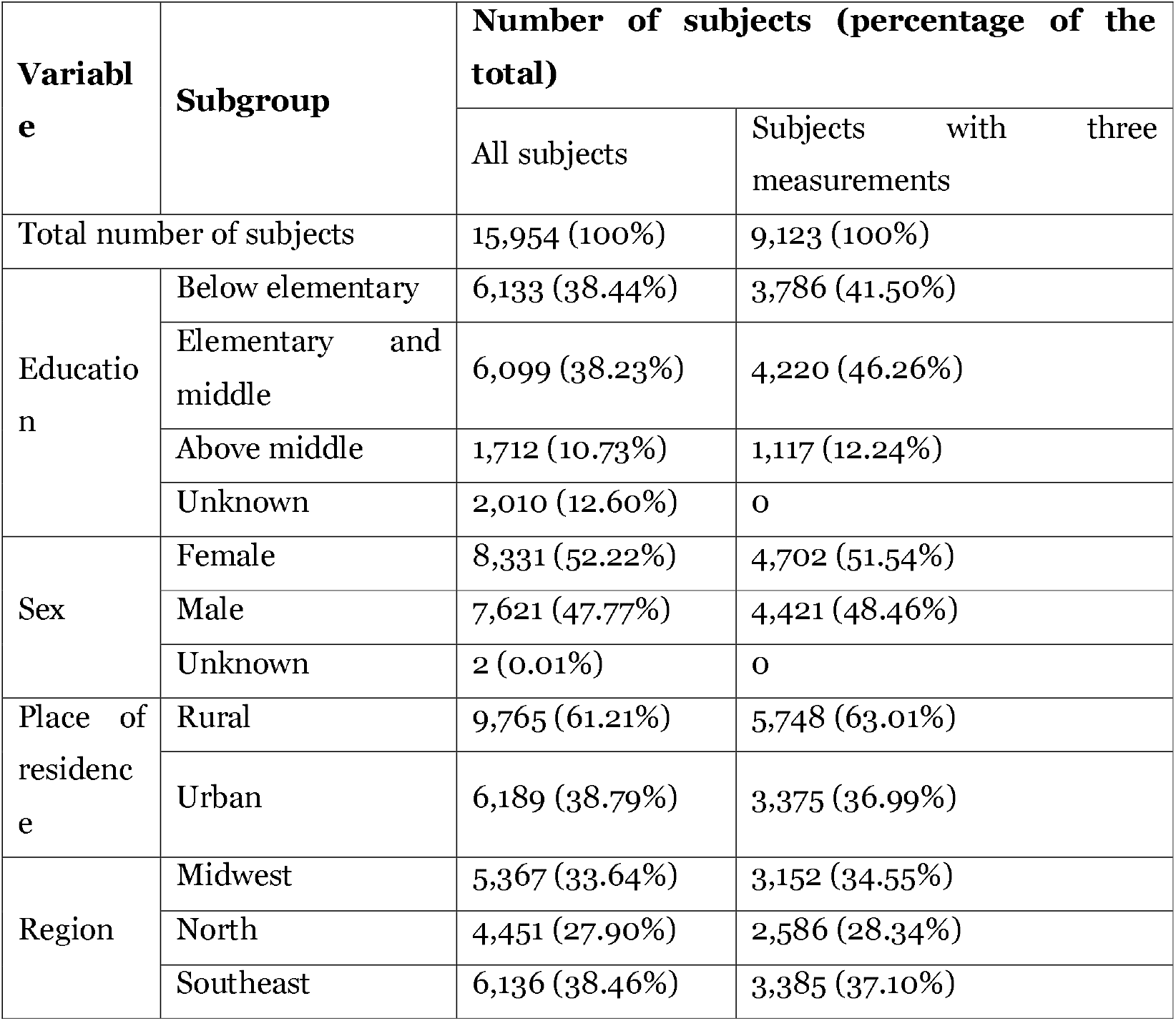
Constant variables of the studied subjects.

| Variable | Subgroup | Number of subjects (percentage of the total) |  |
| --- | --- | --- | --- |
|  |  | All subjects | Subjects with three measurements |
| Total number of subjects |  | 15,954 (100%) | 9,123 (100%) |
| Education | Below elementary | 6,133 (38.44%) | 3,786 (41.50%) |
|  | Elementary and middle | 6,099 (38.23%) | 4,220 (46.26%) |
|  | Above middle | 1,712 (10.73%) | 1,117 (12.24%) |
|  | Unknown | 2,010 (12.60%) | 0 |
| Sex | Female | 8,331 (52.22%) | 4,702 (51.54%) |
|  | Male | 7,621 (47.77%) | 4,421 (48.46%) |
|  | Unknown | 2 (0.01%) | 0 |
| Place of residence | Rural | 9,765 (61.21%) | 5,748 (63.01%) |
|  | Urban | 6,189 (38.79%) | 3,375 (36.99%) |
| Region | Midwest | 5,367 (33.64%) | 3,152 (34.55%) |
|  | North | 4,451 (27.90%) | 2,586 (28.34%) |
|  | Southeast | 6,136 (38.46%) | 3,385 (37.10%) |

**Table 2.**
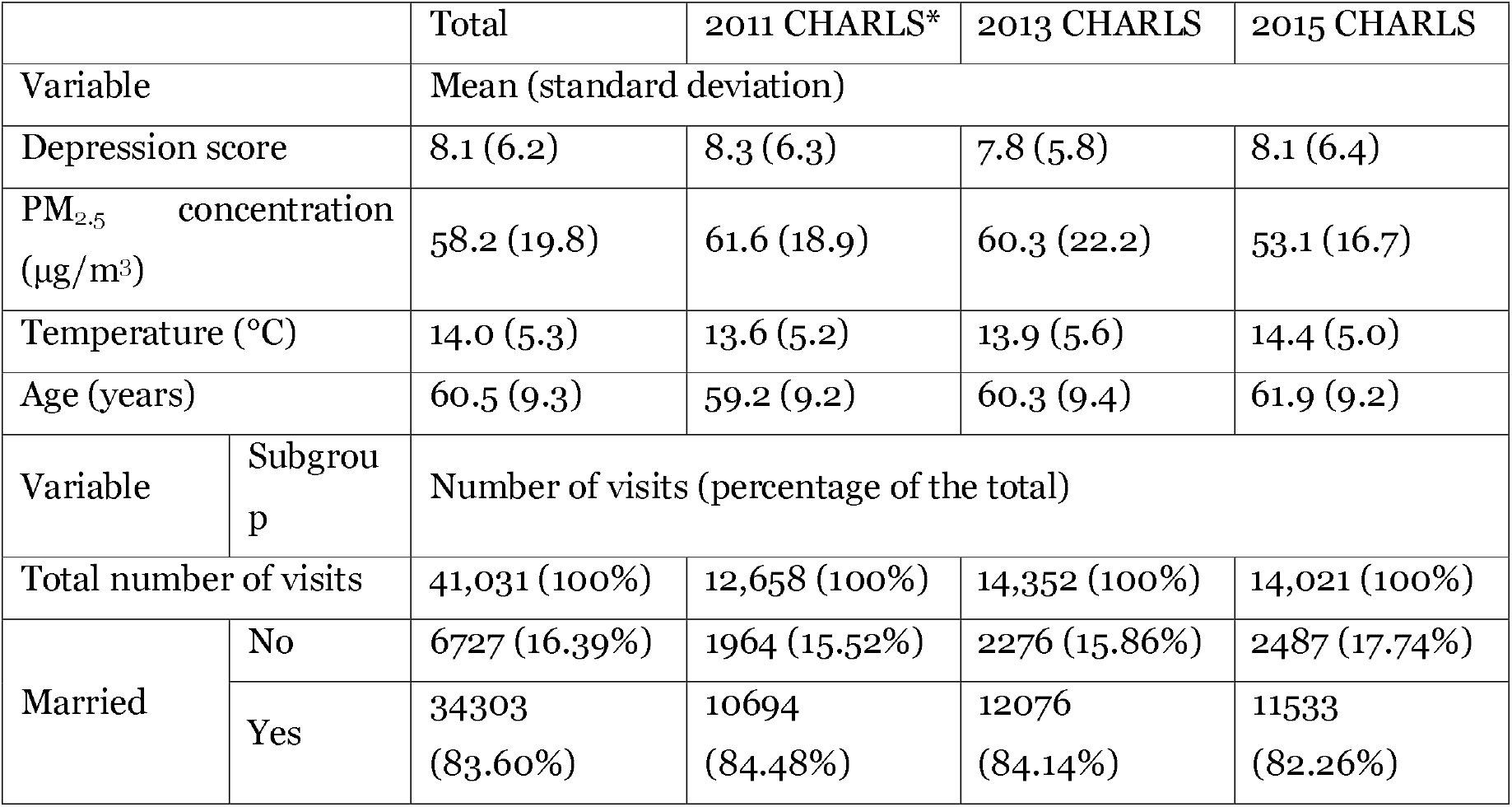
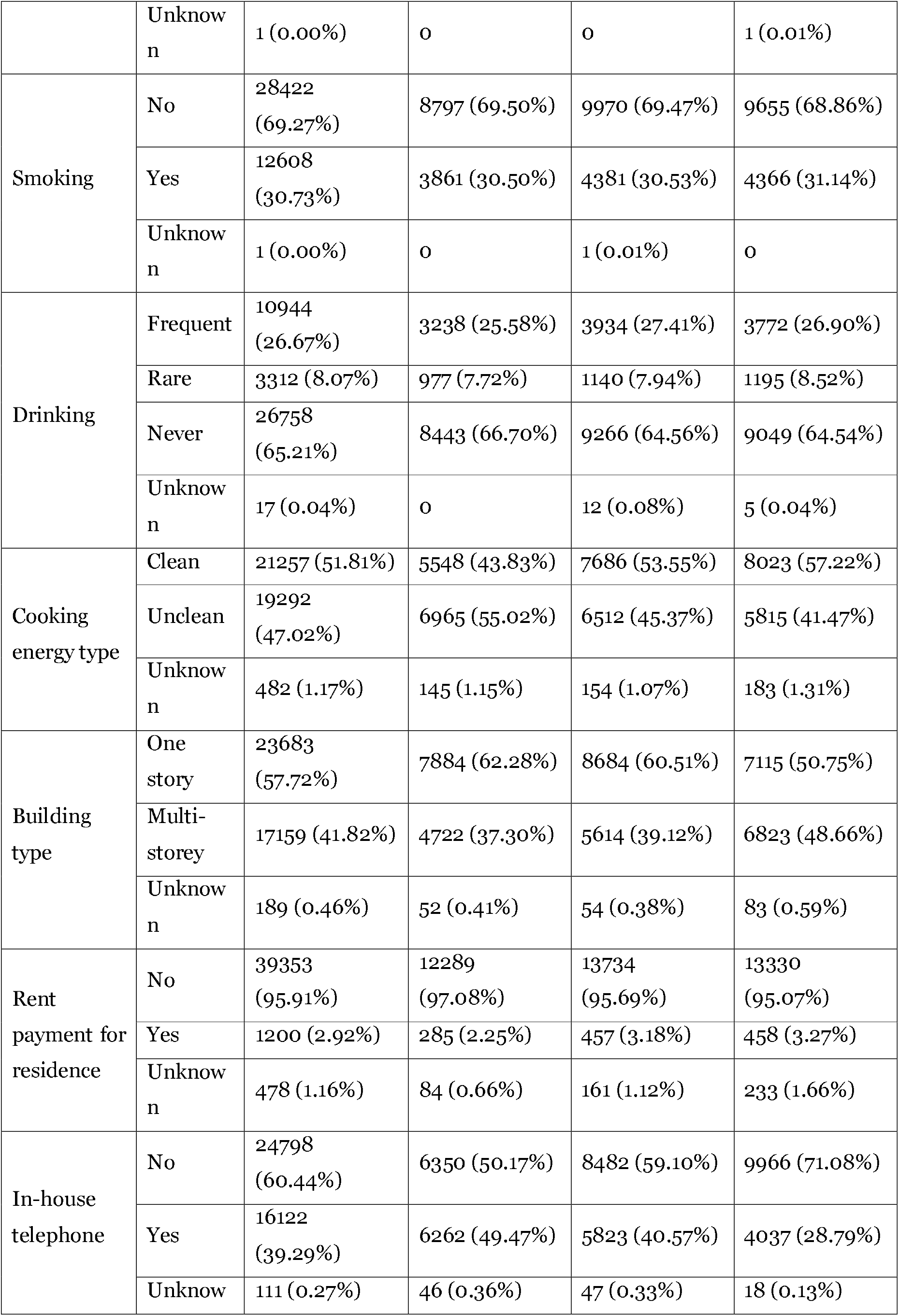
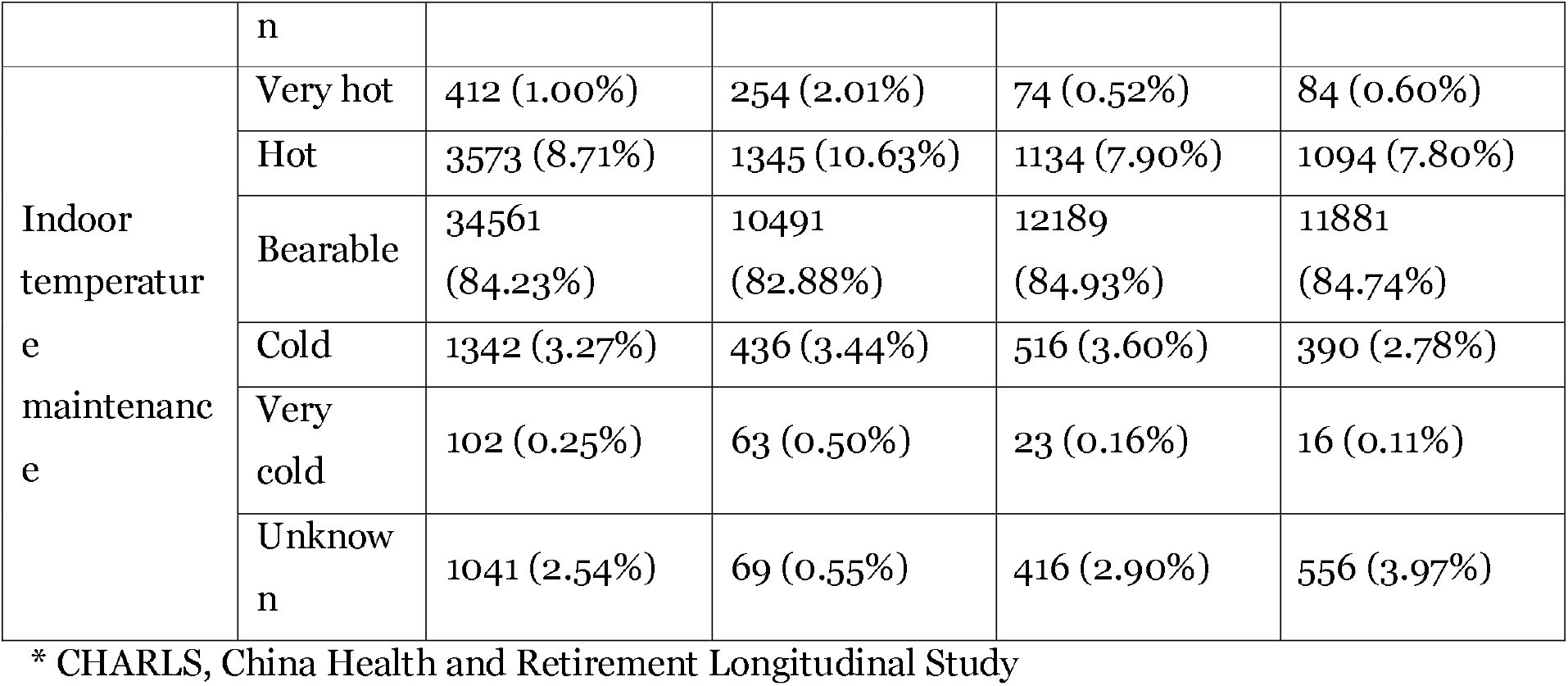
Longitudinal variables of the studied subjects.

### Effect of clean air policy on depression

A DID model of the 10,725 adults who participated in CHARLS 2011 and 2015 showed that the **PM**_2.5_ reduction after CAP was associated with decreased risk of depression. Figure 2 presents the estimated effect of **PM**_2.5_ on the CES-D-10 score, with the adjusted covariates of the DID models. The results of the different models were statistically comparable, considering uncertainties. The estimates consistently suggested that long-term exposure to **PM**_2.5_ was significantly related to the CES-D-10 score. Based on the estimated ER from the standard model (Figure 2), a 10 μg/m^3^ reduction in **PM**_2.5_ concentration was associated with a 4.14% (95% confidence interval [CI]: 0.41–8.00%) decrease in the CES-D-10 score. In sensitivity analyses of DID model, the results were not significantly changed if we changed the settings to control for the effect of clustering of the samples, or did not weigh the samples according to the probability of the reduction in **PM**_2.5_ (Figure S3). However, the estimated effects in the unweighted models were slightly weaker than the weighted results. For instance, after removing the inverse probability weights, the standard DID model yielded an ER of 2.89% (95% [CI]: -0.59%, 6.48%). Because the samples were not optimally randomized (Figure S4), the unweighted estimates (Figure S3) could be slightly biased. To improve the interpretation of the DID model, we also conducted a preliminary analysis based on a binary **PM**2.5 variable, which is documented in supplemental text (S4).

**Figure 2.**
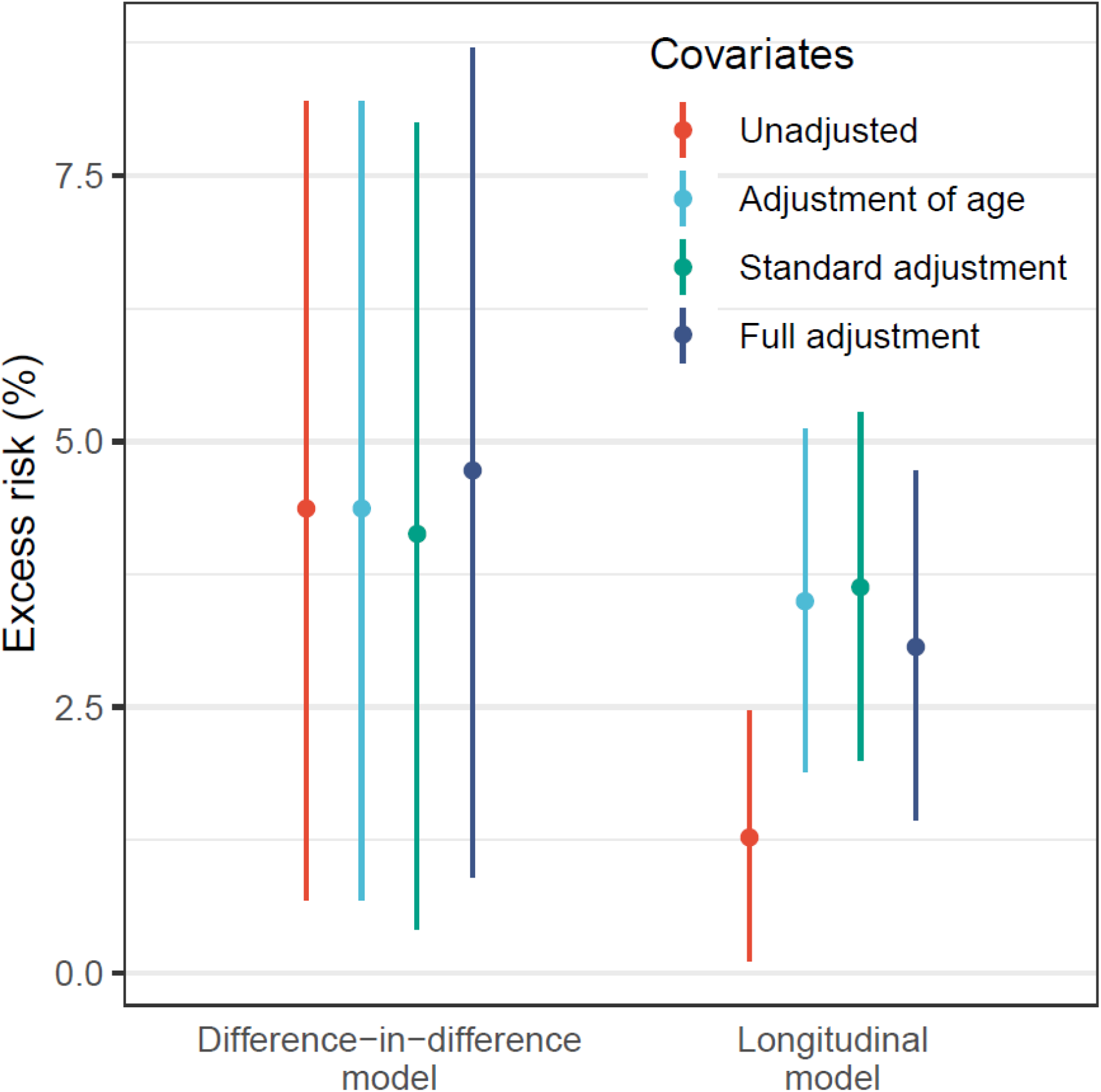
Estimated associations between the depression score and PM_2.5_ concentration. In the difference-in-difference (DID) analysis, the model adjusted for age incorporated age at 2011 as a covariate; the standard adjustment also involved the fixed variables of urban/rural residence, sex, and education, as well as the longitudinal variables of ambient temperature, marriage status, smoking, and drinking. The full adjustment also considered longitudinal changes in cooking energy type, building type, residential rent payment, presence of an in-house telephone, and indoor temperature maintenance. The longitudinal models adjusted for the constant variables in a similar manner as in the DID models; however, adjustment of the longitudinal variables was based on the values recorded in each survey wave, rather than their between-wave changes therein.

### Longitudinal association between PM_2.5_ and depression score

Based on the 41,031 samples from the three CHARLS waves, we quantified the association between **PM**_2.5_ and CES-D-10 score using a longitudinal model (Figure 2). Although the point-estimates from the longitudinal models were slightly lower than those from the DID models, the two types of models reported comparable effects of **PM**_2.5_ on the CES-D-10 score. Additionally, because the longitudinal models incorporated more samples, they had narrower CIs than the DID models. Adjusting for confounders, except age, did not significantly change the longitudinal results. As the studied population ages, a model that does not control for the effect of aging could yield a biased result. According to the model with standard adjustments, a 10 μg/m^3^ change in **PM**_2.5_ concentration was positive associated with a 3.63% (95% CI: 2.00–5.27%) change in the CES-D-10 score (Figure 2 and Table S2). The results of the longitudinal models are listed in Table S2.

In sensitivity analyses, we showed that the association between **PM**_2.5_ concentration and the CES-D-10 score did not vary significantly between sub-populations (Figure S5). The sub-region analysis, which displayed a weak association in the north but strong associations in the Midwest and southeast (P = 0.04; likelihood ratio test). The weak association may have been caused by the small sample size in the north (Table 1) or a higher fraction of natural particles (e.g., dust in the northwest), which are known to be less toxic than anthropogenic particles. The nonlinear analysis showed an approximately linear exposure-response function of **PM**_2.5_ concentration without a no-effect threshold (Figure 3a), which was consistent with estimates from the other models (Table S2). Additionally, we examined the interaction between **PM**_2.5_ concentration and age, and found that the effect of **PM**_2.5_ concentration was weaker in older adults (Table S2). Furthermore, the estimated effect of **PM**_2.5_ was not affected by use of random-or fixed-effects models (Table S3). However, the mixed-effects model, which could be more efficient, but also more biased, than the fixed-effects model ^29^ was used only to enhance the interpretability of the estimates. Finally, the bootstrap analysis (Figure S6) indicated the measurement error caused by using city-level **PM**_2.5_ didn’t significantly change the estimated association. Within a city, where air quality was affected by the same emission-control polices, the temporal trends in **PM**_2.5_ could be similar at different locations. We only ignored 11.75% of total variance in **PM**_2.5_ reductions by using the city-level values (Table S5). Therefore, the measurement error might not undermine our main findings. However, according to the bootstrap analysis, the estimated association became slightly weaker (*i*.*e*., a smaller point-estimate with a larger variance) after correcting for the measurement error, which suggested the effect of **PM**_2.5_ might be underestimated.

**Figure 3.**
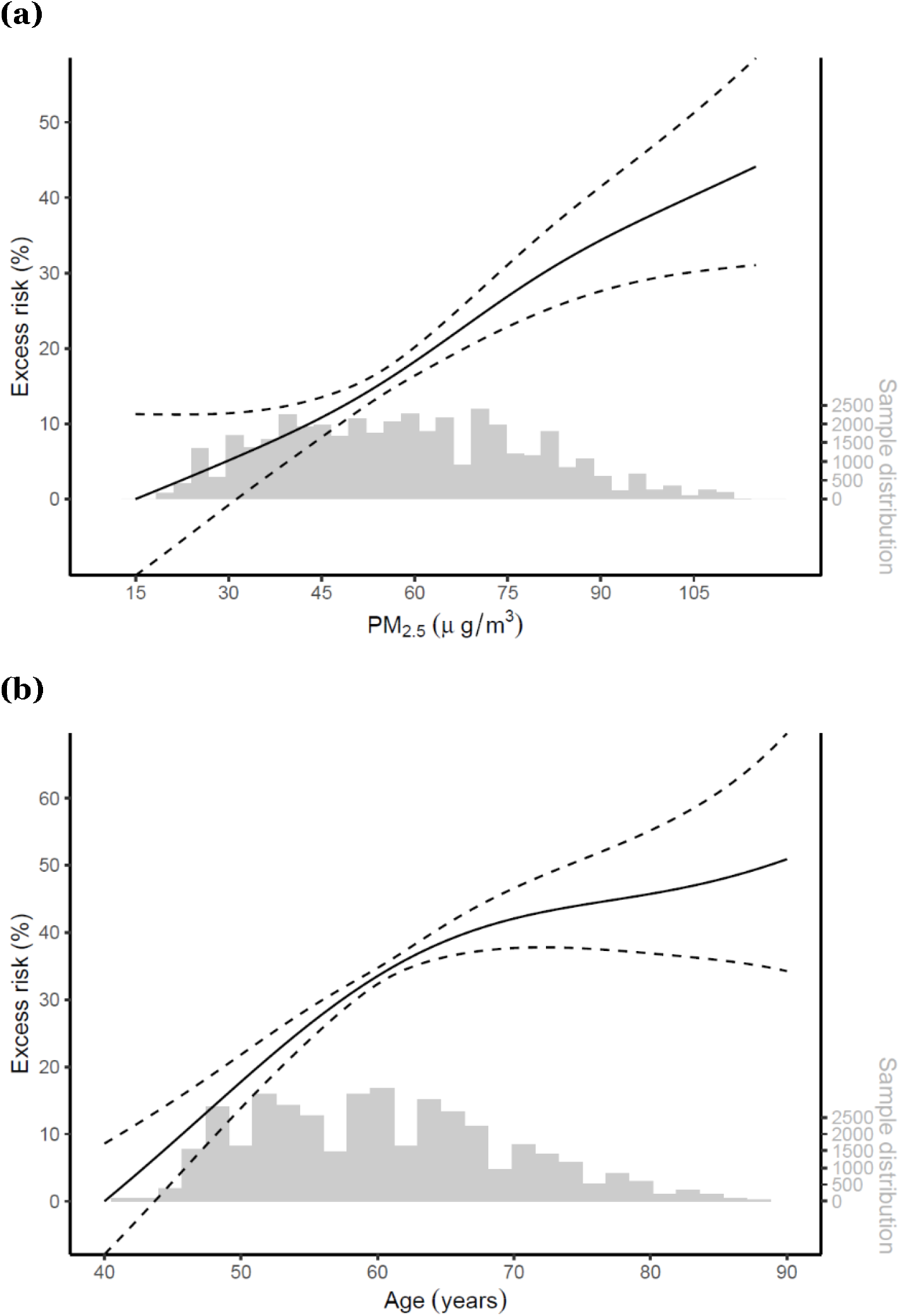
Nonlinear effects of (a) PM_2.5_ concentration and (b) age on the CES-D-10 score. Dashed lines, pointwise 95% confidence intervals. The histograms show the distribution of **PM**_2.5_ exposure and age.

### Evaluating the impact of PM_2.5_

A comparison between the effect of **PM**_2.5_ concentration and that of age showed that they were statistically comparable (Figure 3b). Based on the standard model, the effect of 1 year was of equivalent magnitude to the effect of a **PM**_2.5_ concentration increase of 2.1 μg/m^3^ (95% CI: 1.1–4.2 μg/m^3^). More details on the association between aging and the CES-D-10 score are documented in supplemental text (S5).

The impact of a risk factor on the CES-D-10 score is determined by its effect magnitude and temporal changes. Based on the 9,123 adults who participated in all three waves of the CHARLS, we quantified the impacts of air quality improvement and aging on their mental health status from 2011 to 2015 (Figure 4). According to the results, aging and **PM**_2.5_ reduction were the major drivers of changes in CES-D-10 score. Using the CES-D-10 score obtained before implementation of the CAP (by CHARLS 2011) as the reference, we found that a decreased concentration of **PM**_2.5_ resulted in a relative reduction in the score of 0.27% (95% CI: 0.15–0.40%) and 2.87% (95% CI: 1.61–4.21%) in 2013 and 2015, respectively; in contrast, aging resulted in a relative increase of 1.53% (95% CI: 0.87–2.13%) and 3.08% (95% CI: 1.74–4.31%), respectively (Figure 4). Analysis of the combined impact of age and **PM**_2.5_ (including their interactions), which together resulted in a small reduction in the CES-D-10 score of 0.25% (95% CI: -0.78–1.30%) from 2011 to 2015, showed that the benefit of reducing the **PM**_2.5_ concentration offset the negative effect of aging. Because the impact evaluation was based on a fixed population, we quantified the impact of individual-level aging, which was faster than the aging of the population (Figure S1).

**Figure 4.**
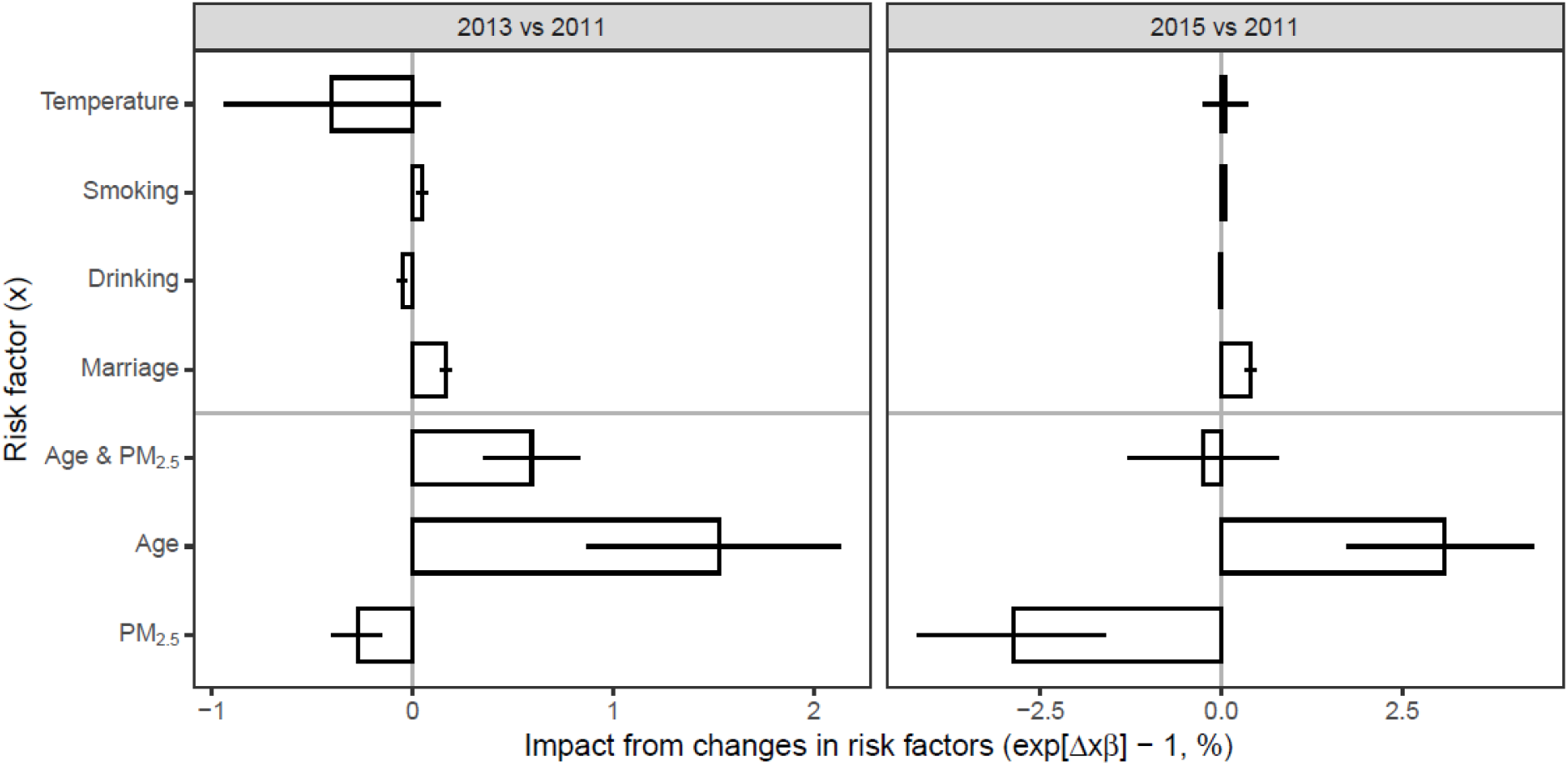
Impacts of reducing PM_2.5_ concentration, aging, and changes in other factors on the CES-D-10 score. The changes in risk factors (Δ*x*) were evaluated using a fixed group of subjects who participated in all three China Health and Retirement Longitudinal Study (CHARLS) waves; the corresponding coefficients were estimated from the longitudinal model with standard adjustment (Figure 2) and presented in Table S3; for each variable, the value in 2011 was acted the reference to evaluate the impact attributable to its temporal changes.

## Discussion

Based on the CHARLS surveys, we found a robust exposure-response function between air pollution and depression, during a quasi-experimental scenario under CAP. Our results also indicate that the effect of **PM**_2.5_ concentration reduction was comparable with that of aging. Although aging partially offset the benefits from reducing **PM**_2.5_ concentrations, the rapidly improved air quality since 2013 could still bring a net positive impact.

An increasing number of studies have reported on the association between air pollution and mental disorders, including depression,^8-10,36-42^ cognitive functions,^6,7,43^ and other indicators of mental health.^5,44-49^ For instance, Fan et al. (2020) conducted a meta-analysis of 637,297 subjects and reported that a 10 µg/m^3^ increase in **PM**_2.5_ concentration is potentially associated with a 12% (95% CI: -3–29%) ER of depression^50^. They also reported that the pooled estimate was sensitive to a cross-sectional study^51^ (which reported an extremely large ER as 1,818%, 95%CI: 189-12,669%), and was changed to 12% (95% CI: 2–23%) after exclusion. Many of these studies examined the association between air pollution and mental illness among residents in China.^3,6,7,39-41,45,47,49^ However, most of them were cross-sectional studies^39-41,45,49^, which contributed to the low confidence level in the meta-analysis and led to an inclusive association between **PM**_2.5_ exposure and depression. Comparing to them, our quasi-experiment is advantageous in controlling for potential confounders, and thus enriches the high-quality evidences. Detailed discussions on the comparison are documented in a supplemental text (S6).

The central government carried out an action plan of air pollution control and prevention from 2013 to 2017 to tackle severe air pollution in China.^15^ Based on a chemical-transport model analysis,^16^ the CAP respectively led to **PM**_2.5_ concentration reductions from 89.5 μg/m^3^ in 2013 to 58.0 μg/m^3^ in 2017, and thus has been show to protect the public health by reducing cardiorespiratory mortalities associated with **PM**_2.5_.^15^ The present study showed that the CAP can also bring benefits to adult mental health.

The present study also examined the effect of aging on depression and compared it to the effect of **PM**_2.5_ concentration. Such an analysis improved the interpretability of the association between **PM**_2.5_ concentration and the depression and has policy implications. The Chinese population is expected to age rapidly because of a low fertility rate. According to the world population projection for the total population of China (Figure S7) (http://dataexplorer.wittgensteincentre.org/wcde-v2/), the mean age of all Chinese adults ≥ 40 years old will increase from 55 years in 2010 to 62–65 years in 2050. Based on our model (Table S2), population aging could offset the benefits from a 14.9–21.3 μg/m^3^ reduction in **PM**_2.5_ concentration. Assuming that air quality meets the current national standard (annual mean **PM**_2.5_ < 35 μg/m^3^) in 2050, the population-weighted concentration of **PM**_2.5_ would decrease by 26 μg/m^3^ (from 61 μg/m^3^ in 2010 to 35 μg/m^3^ in 2017). Therefore, the positive impact of **PM**_2.5_ concentration reduction on depression is comparable to the negative impact of population aging. This rough analysis suggests that the older population requires more stringent air quality standards to protect their mental health. However, notably, the estimated association between age and depression not only reflects the underlying physiological effects (e.g., increased oxidative stress) but also the socioeconomic effects (e.g., decreased income) attributable to aging, and thus it may not be representative of the future population. Therefore, exactly quantifying the combined impacts of a change in air quality and population aging is beyond the capacity of this study. Further prospective studies of the effects of changes in societal and cultural factors on the mental health of an aging population, and air quality, are warranted.

This study was subjected to the following limitations. First, exposure to ambient **PM**_2.5_ was assessed at the city-level due to the lack of specific addresses, which resulted in misclassification of exposure by ignoring the within-city variation in **PM**_2.5_ concentrations. Although such an ignorance might not change the direction of the association between **PM**_2.5_ and depression, it still introduced bias into point-estimate of the association and amplified its uncertainty range (Figure S6). The misclassification might be also caused by falsely specifying the exposure time-window, as well. Because detailed survey dates were unavailable in the open-access CHARLS data, the long-term exposure to **PM**_2.5_ was evaluated using annual means based on monthly scale time-series. Additionally, uncertainties in the **PM**_2.5_ concentration hindcast estimator are another source of exposure misclassification, which usually leads to an underestimated association. To derive an unbiased association between **PM**_2.5_ and depression, advanced assessment techniques, such as personal monitors, should utilized in future studies to reduce exposure misclassifications. Second, the modifiable area unit problem (*e*.*g*., ecological fallacy) due to the city-level aggregation, might also cause bias. Although the subjects screened by the community-based fieldworkers were less likely to change their residential city during the study period, exposure levels of some individuals (*e*.*g*., the residents near city boundaries) might be different from the mean level. Urban and rural residents might have different degrees of modifiable-area-unit problem, but their effect estimates were similar (Figure S5), which suggests a weak impact on our results from the issue. Third, although this study controlled for temporally invariant confounders (e.g., bullying victimization and childhood sexual abuse) and a few longitudinal covariates (including marriage, aging, and alcohol drinking), it may have missed some risk factors for depression. Although the estimates are unlikely to be confounded by some of them (e.g., drug abuse), failure to adjust for all potential confounders could have biased our results. Fourth, this study utilized **PM**_2.5_ mass concentration as a general indicator of ambient air quality, which may have underestimated the complexities of air pollution toxicity. For instance, **PM**_2.5_ is a mixture of particles of different chemical species and sizes, and between-component variations in toxicity have been shown for **PM**_2.5_ (Han and Zhu, 2015, Han et al., 2016). Additionally, the estimated association between **PM**_2.5_ and depression might not be attributable to only itself, but also to other air pollutants, such as ozone,^52^ that co-vary with **PM**_2.5_. Therefore, further studies on the effects on mental health of other air pollutants are warranted. Finally, some CHARLS subjects dropped out of the study for various reasons, which may have biased the results.^53^ Also, the limited information on missing data from the publicly available CHARLS datasets hampered the derivation of sampling weights for the statistical models, reducing the national representativeness of the findings. Although the analysed samples were comparable to the baseline representative population recruited in CHARLS 2011, there were still slightly differences in some aspects, such as the geographic distribution (Figure S9). Given the potential heterogeneity in the association between **PM**_2.5_ and depression (Figure S5), the estimates from a less representative sample might be biased away from the average effect of target population. Given the above limitations, the causality of our findings should be interpreted cautiously.

In conclusion, we found a robust association between an increase in **PM**_2.5_ concentration and depression risk in a nationwide sample of adults in China. Due to CAP, the exposure concentrations of **PM**2.5 were decreased rapidly, which subsequently improved mental health. This study not only enriched the epidemiological evidence on the adverse effects of air pollution on mental health, but also indicated implementing CAP could improve mental wellbeing of adults.

## Data Availability

The study is based on public data.

## Contributor

T.X. & T.Z. designed this study; T.X., Y.Z., G.G. & Q.Z. prepared and analysed the data; T.X., T.G. & T.Z. drafted the manuscript; Y.Y. & T.X. designed and performed the bootstrap analysis; all co-authors revised the manuscript together.

## Acknowledgements

Thanks to the China Center for Economic Research, National School of Development, Peking University for providing the CHARLS data. This work was supported by National Natural Science Foundation of China (41701591, 81571130100, 81903392, and 41421064), Ministry of Science and Technology of China (2015CB553401 and 2018YFC2000400) and Energy Foundation (G-1811-28843). The funders had no role in study design, data collection and analysis, decision to publish or preparation of the manuscript.

## Date sharing statement

This study is based on publicly-available datasets. The health data can be accessed from the website: http://opendata.pku.edu.cn/, and the environmental data can be accessed from the websites: http://www.meicmodel.org/dataset-phd.html.

*Editor note: The Lancet Group takes a neutral position with respect to territorial claims in published maps and institutional affiliations*.

