## Supplemental materials for "Long-term PM_2.5_ exposure and depressive symptoms in China: a quasi-experimental study"

**Supplementary Text**

**S1 Difference-in-difference analyses**

In quasi-experiments, the difference-in-difference (DID) approach has been typically utilized to evaluate the impact of policy interventions,^1^ including in studies of the effect of air pollutants on health.^2-4^ We utilized the DID model to examine the association between PM_2.5_ concentration reductions on the CES-D-10 score change.

The reductions in PM_2.5_ from 2011 to 2015 were driven by the clean air actions;^5^ the magnitude of the changes can be interpreted in the context of different emission-control measures, and might be unrelated to conventional risk factors for depression, such as aging. In conventional DID analysis of a binary exposure, subjects residing in areas subject to more stringent policies (which resulted in a larger PM_2.5_ reduction) were included in the treatment group, and living in areas subject to less stringent policies were the controls (Figure S2). The between-group difference in the change in CES-D-10 score from 2011 (i.e., before CAP) to 2015 (i.e., after CAP) could be taken as a proxy of the effect of policy. Because PM_2.5_ is a continuous variable, a regression analysis of the association between changes in PM_2.5_ and CES-D-10 score was performed. The DID analysis requires treatments and controls to be randomly assigned. Therefore, we utilized the inverse probability of changes in PM_2.5_ as the regression weights. The probability was calculated using a Gaussian regression model of PM_2.5_ against the covariates, including residence (urban/rural), sex, educational level, age at 2013, and temperature (Supplemental codes). Because the subjects were clustered by household and sampling site (*i.e.*, community), we also incorporated the corresponding random effects into the regression model. DID analysis is advantageous by controlling for some unmeasured confounders (*e.g.*, aging) by the study design itself.^1^ Because the CHARLS 2013 samples were partially influenced by the air quality intervention, the DID model included only 10,725 samples from the 2011 and 2015 waves (Figure S1). Because the distribution of the absolute changes in CES-D-10 score was skewed and leptokurtic, we modeled the relative changes. To do this, we added 1 to the original CES-D-10 score and calculated the changes on a logarithmic scale such that the skewness of the distribution was reduced (skewness = 0.03), as was the leptokurtosis (kurtosis = 0.65). We calculated the excess risk (ER) for each 10 μg/m^3^ change in PM_2.5_ concentration using the following equation: ER = [exp(10β) − 1] × 100%, where β denotes the estimated regression coefficient.

In sensitivity analyses of the DID model, we first examined whether our results were altered by adjustment for covariates. The covariates included constant characteristics (i.e., urban/rural residency, sex, educational level, and age in 2011) and changes in longitudinal variables (i.e., ambient temperature, marriage status, smoking, drinking, cooking energy type, building type, residential rent payment, presence of an in-house telephone, and indoor temperature maintenance). The missing covariate values were first imputed using the chained-equation approach. ^6^ Next, we presented the results of the re-estimated DID models without regression weights, and evaluated the importance of randomly assigning the samples across different levels of exposure change. Additionally, we removed the random effects to control for spatial clustering of the samples from the original DID models or changed them through replacing the community term by the city term, and recalculated the effect estimates. Finally, since the DID model presumed the outcome variable processed in the same pattern between treatments and controls, we examined whether the CES-D-10 score variations during the pre-treatment period (2011-2013) were associated to the treatment-control assignment.

**S2 A bootstrap method to correct the exposure measurement errors**

In the publicly available version of CHARLS datasets, there was no specific address for all the surveyed subjects, in order to protect confidentiality. Because of that, all of our analyses were depended on city-level averages of PM_2.5_ concentrations. Using the averages would introduce exposure measurement errors into our association models. Previous studies found that the error would both introduce bias into the point-estimate and enlarge its variance. To correct measurement errors in exposure, Szpiro et al. derived a bootstrap method ^7^. The well-established method was designed for measurement errors caused by spatial misalignment (*i.e.*, health outcomes were measured at place a, while the exposures were monitored at the different by nearby places b), and can be also utilized here after a simple modification. The basic idea is to take the data-generation mechanism of the exposure assessment into the model estimation, through designing an appropriate bootstrap procedure to derive the empirical distribution for the association estimator (*β*), given the measurement error. The bootstrap method can specified as follows:

1. Estimate the association (*β*) between PM_2.5_ and a biomarker using the fully-adjusted regression (Equation 1);
2. Randomly select a location based on the finest-available exposure data (*i.e.*, a 0.1° × 0.1° pixel of the PM_2.5_ map) within the corresponding city as the address for each subject;
3. Assign *pseudo*-*true* exposure values (PM_2.5_^*^) based on the addresses and the gridded monthly maps of PM_2.5_;
4. Simulate *pseudo*-*true* outcomes (*y*^*^) based on Equation 1 and the *pseudo*-*true* exposures (PM_2.5_^*^);
5. Re-estimate the association (*β*^*^) between *pseudo*-*true* outcomes (*y*^*^) and city-level exposures (PM_2.5_);
6. Repeat the steps 2-5 iteratively to mimic the data-generation procedure, which causes the exposure measurement errors, and utilize the sampled distribution of *β*^*^ as the estimation with error correction.

We conducted a bootstrap-based estimation with 500 simulations, and presented the empirical distribution of *β*^*^ in Figure S6.

**S2 Descriptive summary**

Among the 15,954 studied adults, there were more rural adults (61.21%) than urban adults (38.79%), decause of the different response rates.^8^. Although the mean CES-D-10 score in the 2013 (score = 7.8) or 2015 CHARLS (8.1) was smaller than that in the 2011 CHARLS (8.3), the comparison is not indicative of the trend in depression, given the between-wave difference in the surveyed population. The long-term mean PM_2.5_ concentration for 2015 CHARLS subjects was 53.1 μg/m^3^, which was considerably lower than the value for the 2013 (60.3 μg/m^3^) or 2011 subjects (61.6 μg/m^3^). The changes in PM_2.5_ concentrations were within our expectations and were attributed to the clean air actions in China. However, there were many other potential depression risk factors that changed in opposite directions during the study period. For example, we observed an increase in the fraction of unmarried adults from 15.52% in 2011 to 15.86% in 2013, and then to 17.74% in 2015. Marriage results in mental health benefits;^9^ thus, this trend increased the CES-D-10 score. Living conditions, such as in-house bathing facilities and the ability to maintain the indoor temperature, have improved during the study period, which may have reduced the CES-D-10 score. Additional information on the longitudinal variables can be found in Table 2. Because of the complexity of the drivers of depression, the lack of similarity in the trends of CES-D-10 score and PM_2.5_ should not be interpreted as evidence against our hypothesis.

**S3 Results of Preliminary difference-in-difference model**

Among the 10,725 adults who participated in CHARLS 2011 and 2015, 741 had an increased concentration of PM_2.5_ after the CAP. Most of them lived in the places with low or moderate level of PM_2.5_ pollution (Figure S2). To illustrate the DID methodology, we conducted a preliminary analysis of binary PM_2.5_ exposure (Figure S2), and set those subjects as controls (i.e., were not affected by the policy) and the others as the treatments. From 2011 to 2015, the mean CES-D-10 score of the controls increased by 3.7%, but that of the treatment group decreased by 0.9%. The between-group difference in the CES-D-10 score change was significant (P = 0.026), likely due to the intervention. In contrast, between-group difference before the completely implying the actions (2011-2013) was not statistically significant (P = 0.48; Table S1). The results (Table S1) suggest that the treatments and controls had a similar pattern of CES-D-10 score before the intervention.

**S4 The estimated association between age and depression score**

This study also examined the associations between age and the CES-D-10 score, so as to better interpret the mental health effects of PM_2.5_ concentrations. The standard model indicated that the CES-D-10 score increased by 0.76% (95% CI: 0.45–1.07%) for an adult who aged by 1 year (Table S2). The association was robust, given different sets of adjusted covariates (Table S2). Subgroup analyses (Figure S5) suggested that the effect of age was not considerably modified by sub-region or sub-population indicators. The nonlinear analysis (Figure 4b) showed a sublinear exposure-response function for age, suggesting a weak marginal effect for adults older than ~70 years. Additionally, considering the collinearity between age and the temporal trend, we re-estimated the effect within each CHARLS wave, based on the cross-sectional comparison of CES-D-10 score among adults of different ages. The results of the cross-sectional analysis (Table S4) were consistent with the estimates from the longitudinal models (Table S2).

The analysis of the interaction between age and PM_2.5_ suggested that the PM_2.5_-depression association is weaker among older adults (Table S2). Although few similar depression findings have been reported, the relative risk of PM_2.5_ is assumed to decline with age for the exposure-response functions of other outcomes, such as mortality.^10^ Because the risks factors other than air pollution increase with age, such an effect-modification is feasible.

**S5 Comparison between the quasi-experimental study and other designs**

Mental health can be affected by a number of complex factors, and comprehensively controlling these factors is critical to minimize bias. However, this may be difficult to achieve in a cross-sectional study. For example, Tian et al. conducted a cross-sectional study of 6,630 older adults (≥60 years old) from the 2013 CHARLS to associate the CES-D-10 score with sulphur dioxide emissions, and reported a U-shaped exposure-response function,^11^ which is biologically implausible and may have been caused by a failure to adjust for potential confounding factors. In the DID models and longitudinal models, the CES-D-10 score of a subject was compared to another observation of the same subject. Thus, the intra-individual comparison controlled for many unmeasured mental health risk factors (e.g., genetics), which varied only inter-individually. Therefore, our longitudinal study is more powerful than the previous cross-sectional analysis.^11^

The quasi-experimental design based on the policy-driven rapid changes in air quality also added power to this study. The intervention exerted a marked effect on PM_2.5_ exposure, which may have had a significant effect on the health outcomes. For instance, Pun, et al. ^12^ conducted a longitudinal study of the two waves of the National Social Life, Health and Aging project conducted in the United States (US), and examined the effect of PM_2.5_ on depression based on an exposure contrast of 2.3 μg/m^3^ from measurements separated by 5 years (annual PM_2.5_ concentration = 11.1 and 8.8 μg/m^3^ for the 2005–2006 and 2010–2011 waves, respectively). Compared to the US study, the mean difference of PM_2.5_ in our study is 8.5 μg/m^3^ during a 5-year period (Table 2), and may have an easily-distinguishable impact on health.

**Supplementary Figures**

(a)

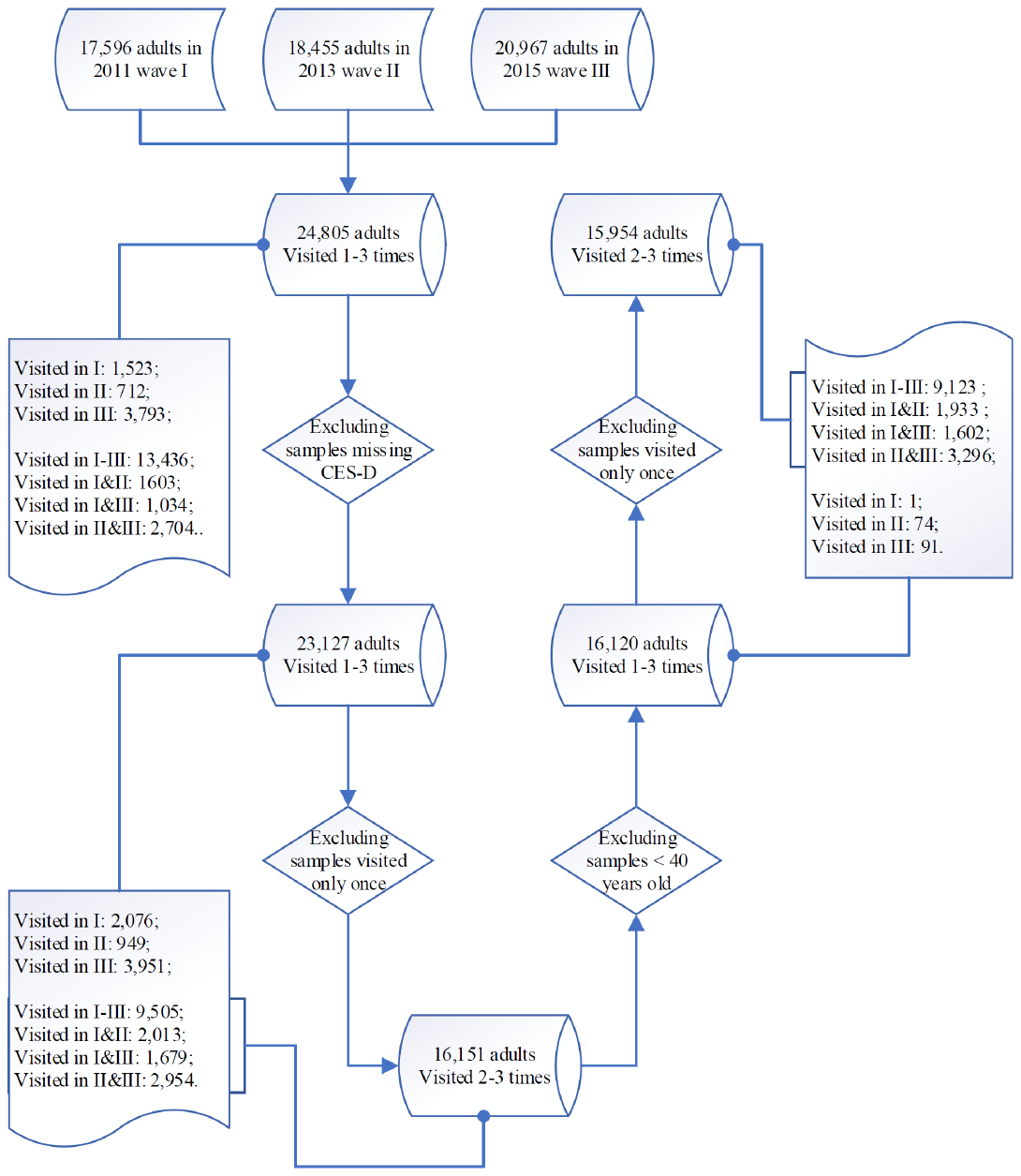

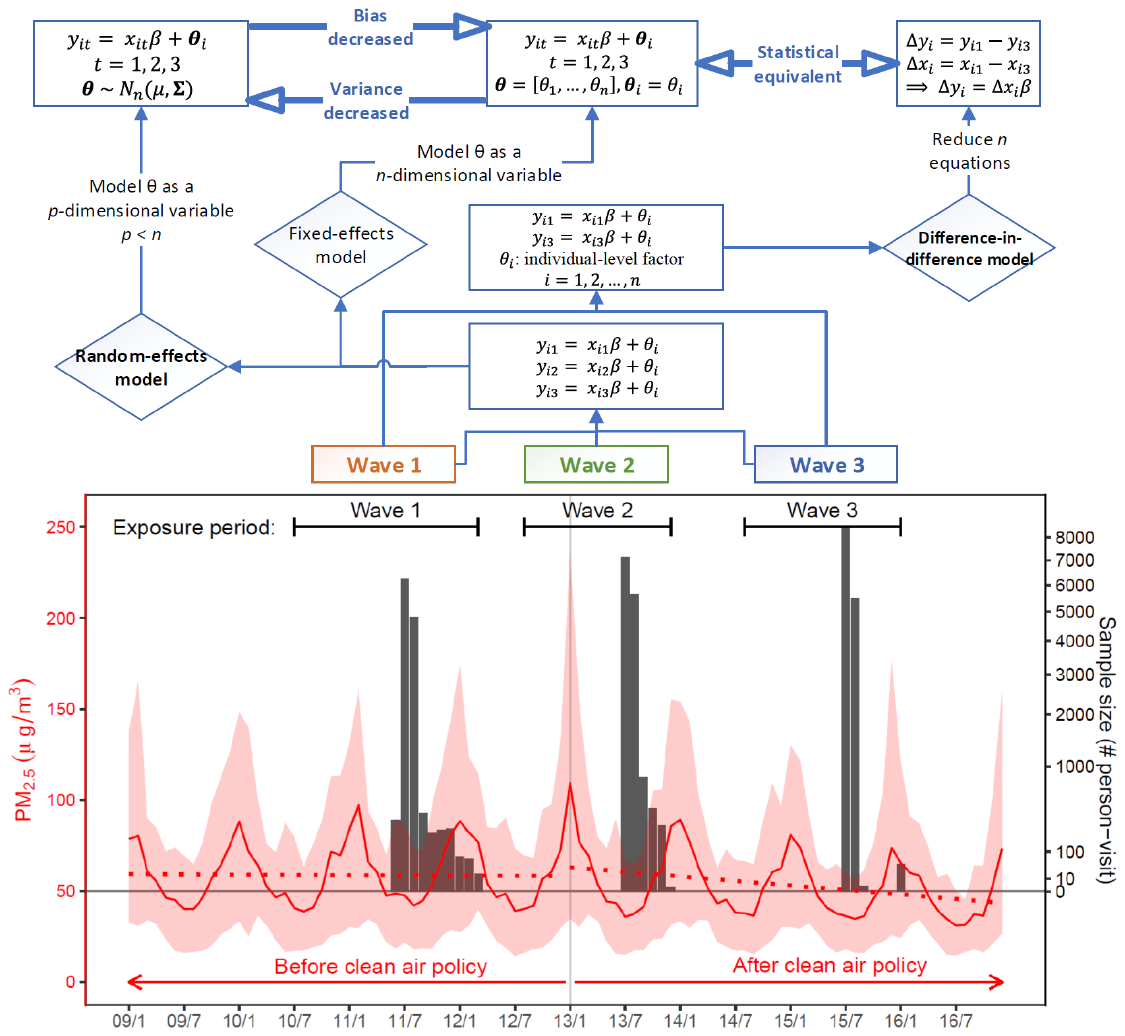
(b)

Figure S1. Study design. Panel (a) presents the data cleaning procedure and the detailed sample sizes. Panel (b) presents ideas underlying the study design and the procedures of data analyses. Bottom of panel (b) presents the study periods covered by the samples (black bars, right y-axis), and monthly PM_2.5_ concentrations (solid red lines, left y-axis), averaged across the sampled cities. The red ribbon presents 95% confidence intervals for the corresponding average PM_2.5_ concentrations; and dashed lines present linear trends in PM_2.5_ before and after the policy intervention. The top of panel (b) presents the data-analysis procedure on how to evaluate the health effect of PM_2.5_ based on within-individual comparisons between waves. The equations are simplified versions of the models that we utilized in data analyses.

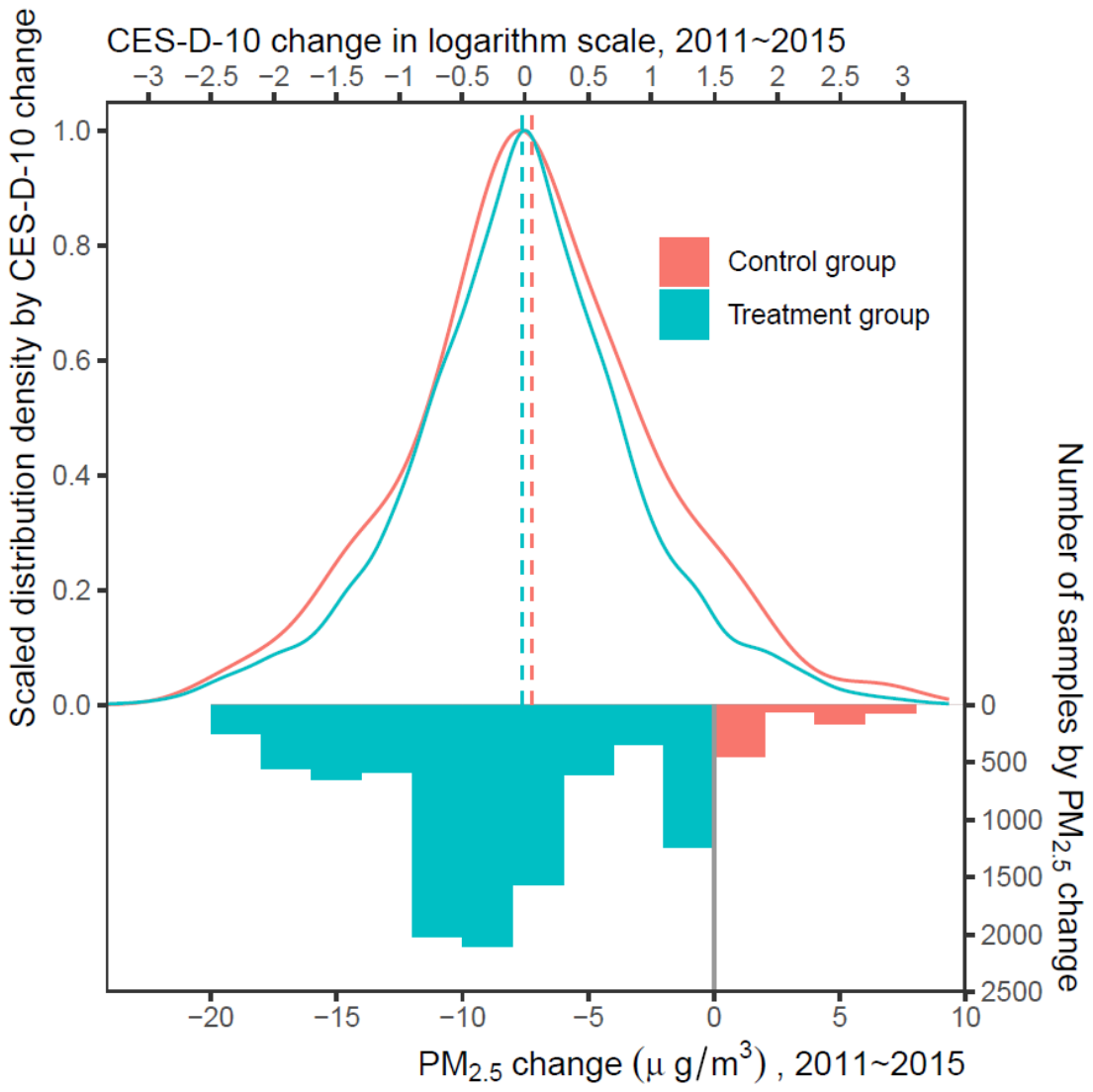
 (a)

(b)

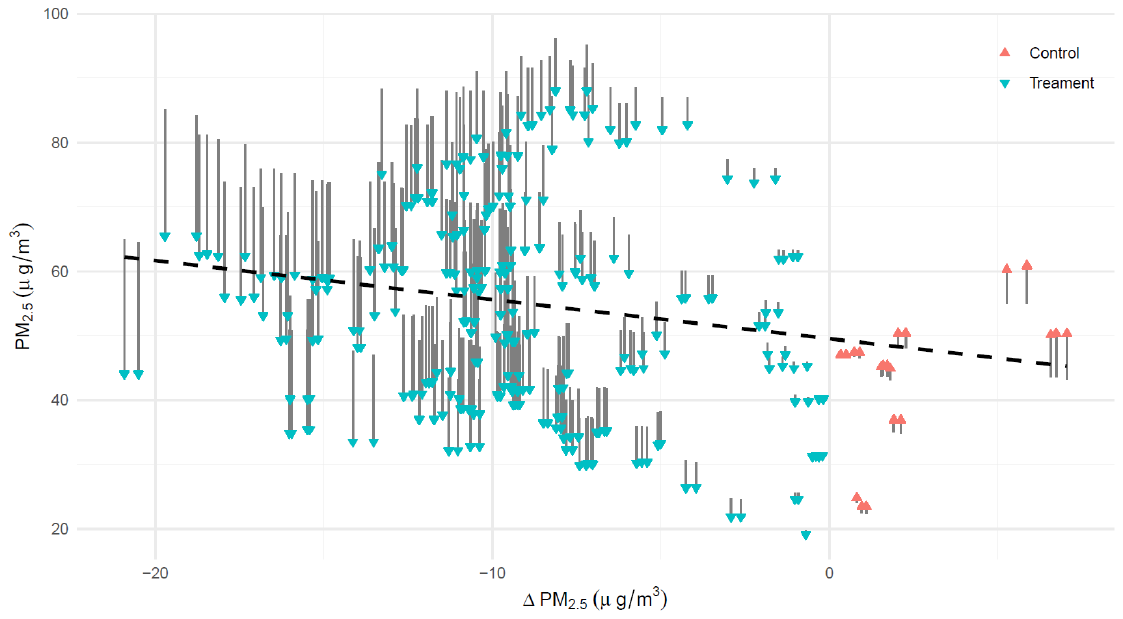

Figure S2. Design of the preliminary difference-in-difference (DD) analysis. In panel (a), the histogram (bottom x-axis and right y-axis) shows the sample distribution according to differences between the PM_2.5_ exposure of CHARLS 2011 and CHARLS 2015. According to the changes in PM_2.5_, the samples were classified as controls (PM_2.5_ change ≥ 0) or treatment (< 0). Solid lines (top x-axis and left y-axis), distribution of CES-D-10 changes (logarithmic scale) among the controls or treatment group; dashed lines, means of the corresponding distributions. Panel (b) shows the detailed changes of PM_2.5_. The bar shows the variations and the triangle shows their directions. The dashed regression line shows the PM_2.5_ changes were weakly and negative correlated with the baseline pollution levels.

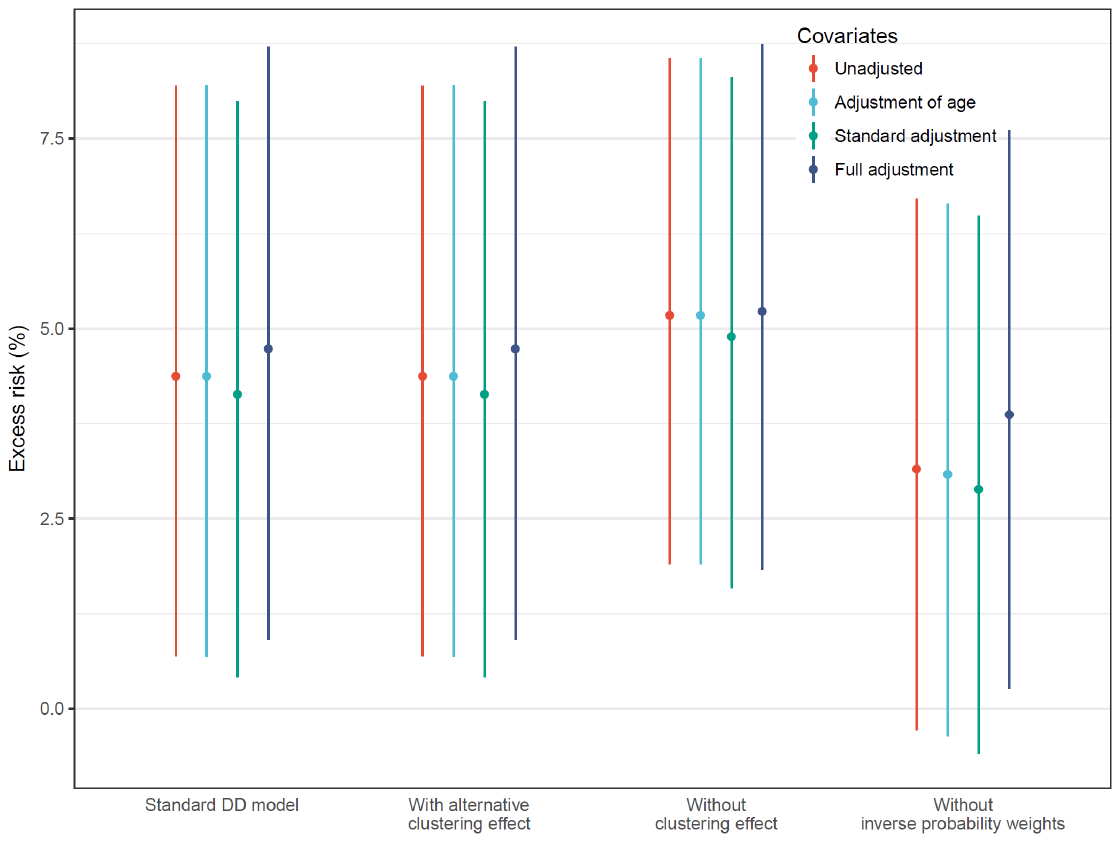

Figure S3. Estimated associations between CES-D-10 and PM_2.5_ concentration derived using the indicated DD models. In the standard model, the clustering effect was modeled at household and community level; in the alternative setting, the clustering effect was at household and city level.

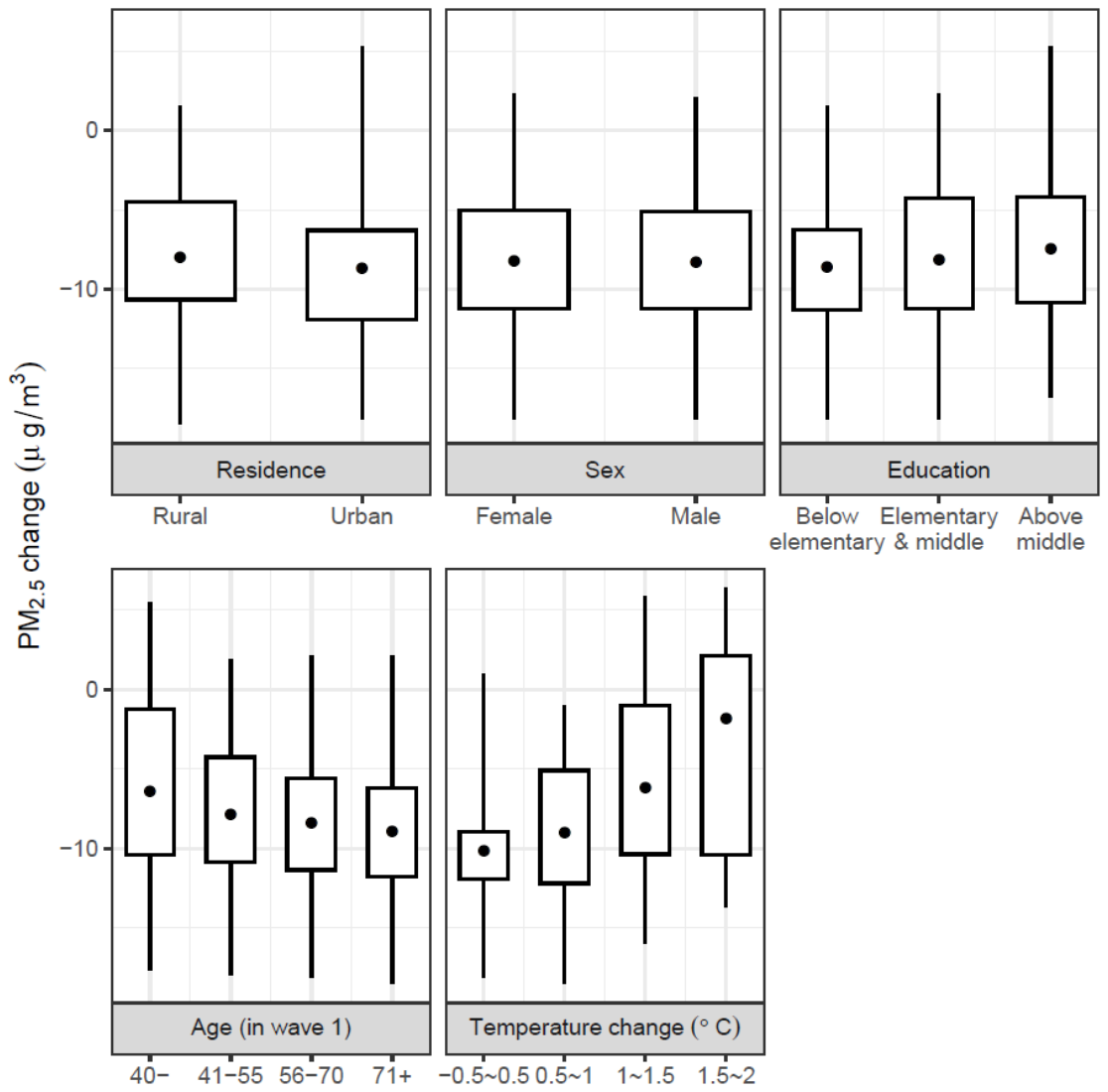

Figure S4. Distribution of changes in PM_2.5_ (from 2011 to 2015) according to the indicated variables.

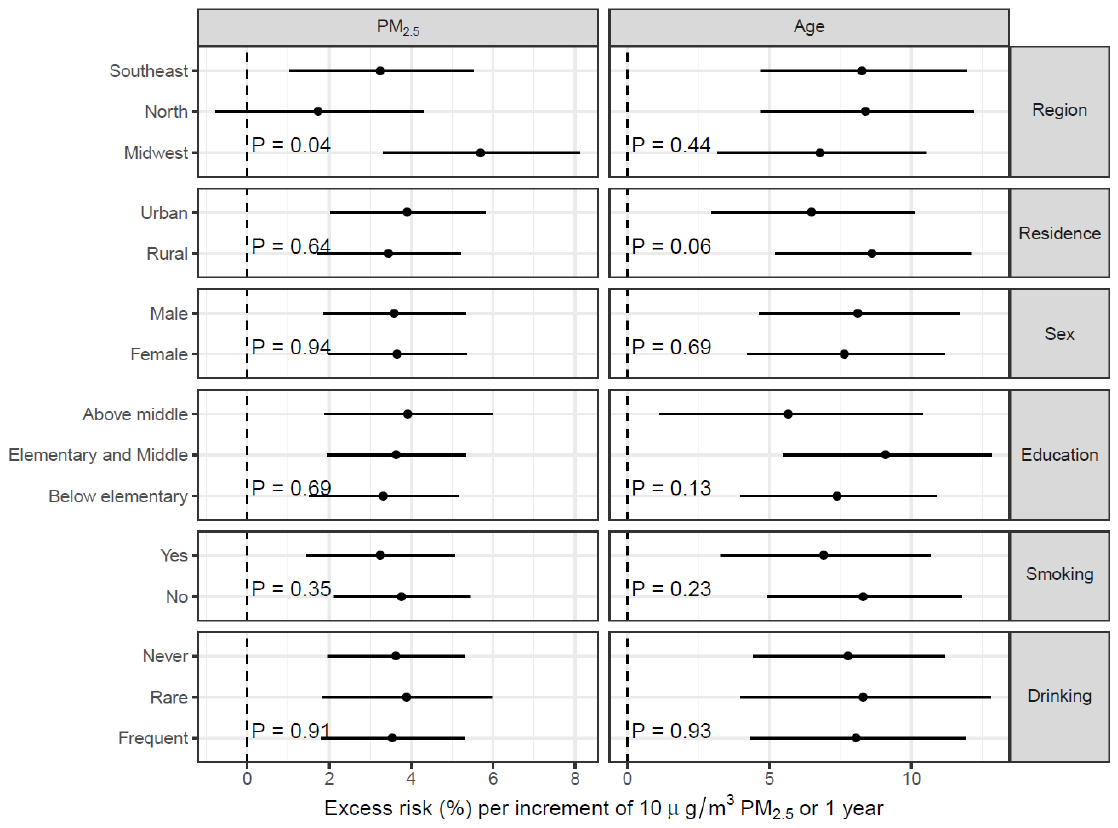

Figure S5. Subgroup-specific associations between CES-D-10 and PM_2.5_ concentration or age. Error bars, 95% confidence intervals. The P-values for tests of the null hypothesis that the point-estimate of the association was identical between subgroups are shown. The associations were estimated using the standard model.

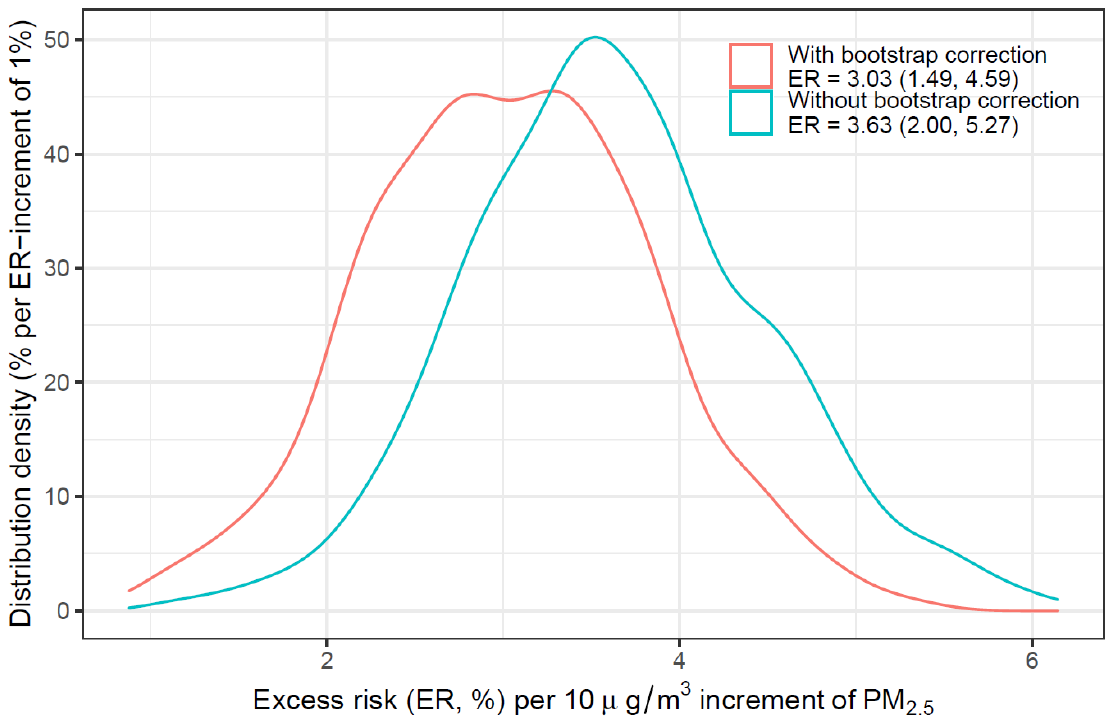

Figure S6 The bootstrapped distribution for the estimated association between PM_2.5_ and depression risk after correcting the measurement errors induced by using the city-level exposures.

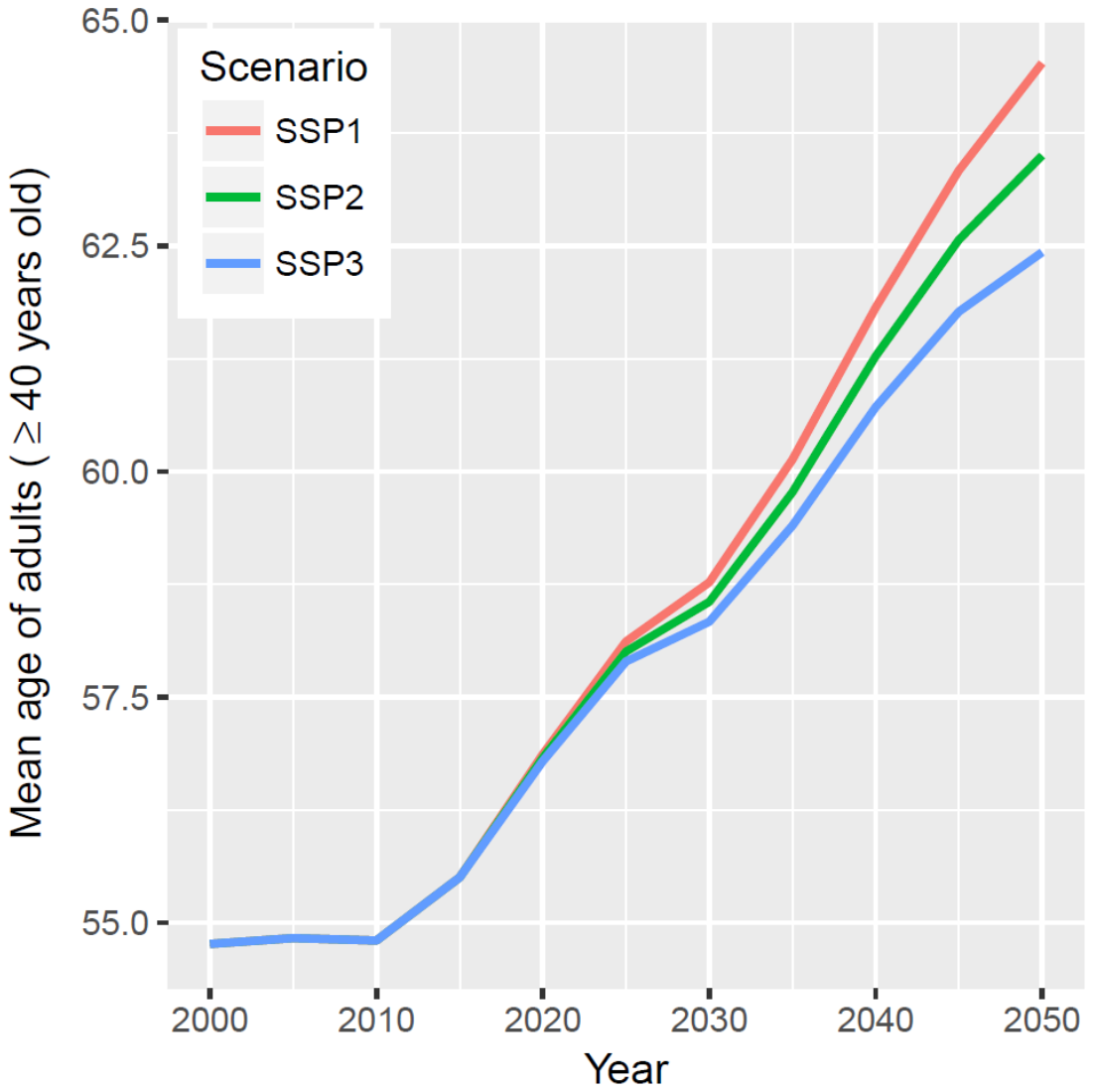

Figure S7. The projected mean age of adults ≥ 40 years old in China under different shared socioeconomic pathways (SSPs). Data source: http://dataexplorer.wittgensteincentre.org/wcde-v2/ (accessed on June 26, 2019).

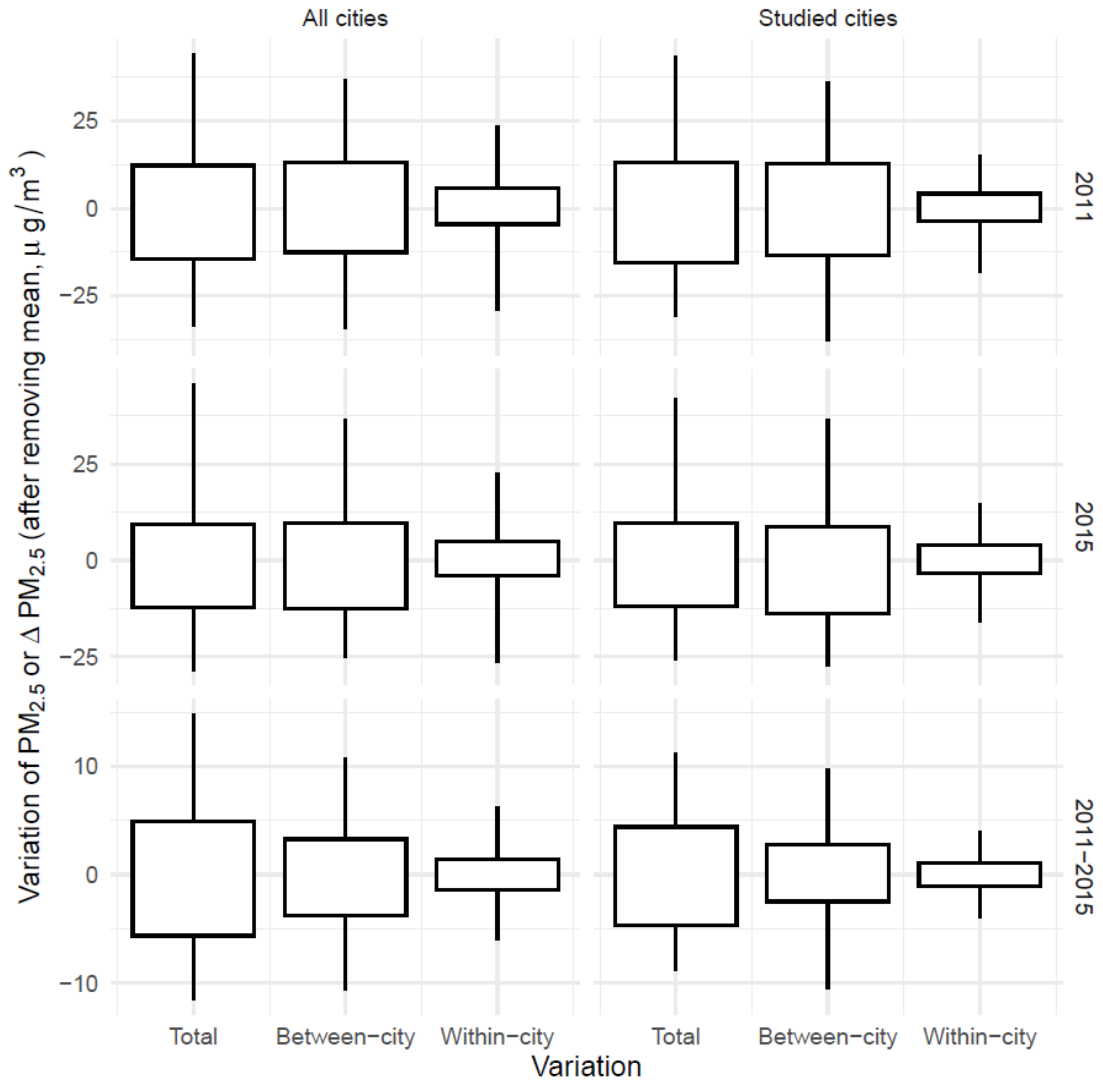

Figure S8 Total, between-city or within-city variation in the gridded annual concentrations of PM_2.5_.

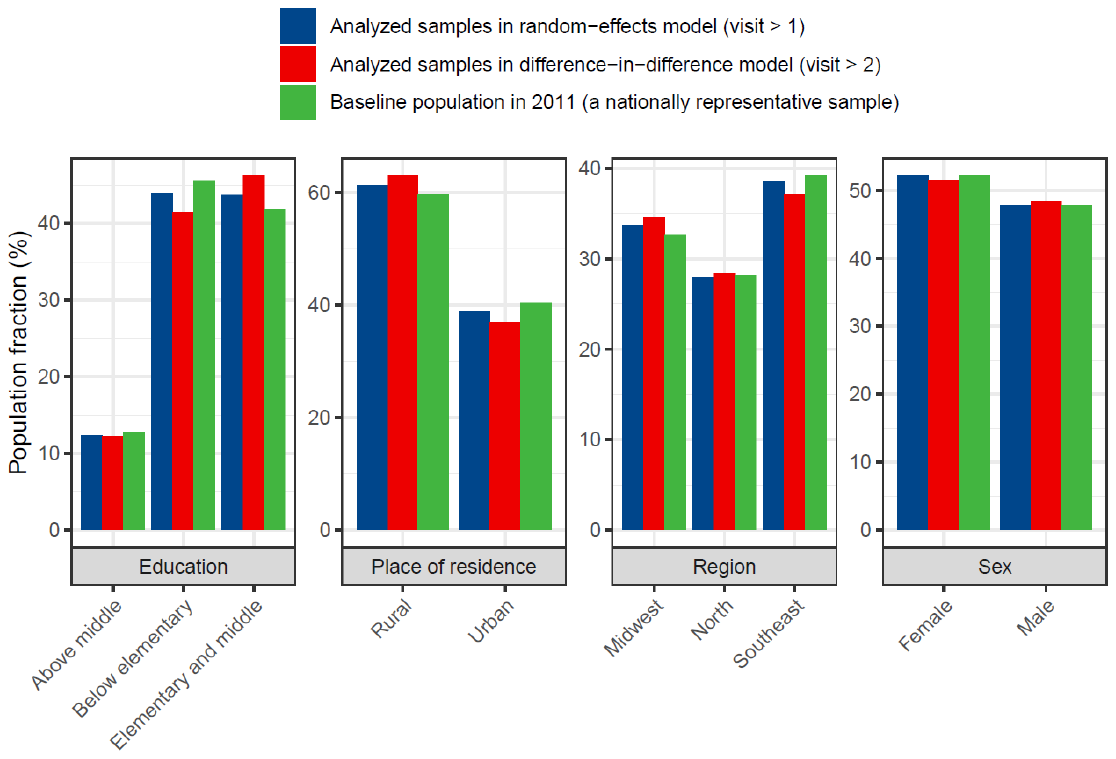

Figure S9 Comparison between the analyzed samples and the CHARLS baseline population (*i.e.*, the 2011 wave), which can be utilized a representative sample of Chinese adults.

**Supplementary tables**

Table S1. Estimated the difference in the mental health change between the controls and treatments (which were defined by the PM_2.5_ reductions due to the clean air actions during 2011-2015) during the pre-treatment period (2011-2013).

| Settings in the difference-in-difference model | | Excess risk (%) with 95% confidence intervals | P-value |
| --- | --- | --- | --- |
| Binary variable  (PM_2.5_ > 0) | Unadjusted | 2.47 (-4.28, 9.69) | 0.482 |
|  | Standard adjustment^*^ | 0.30 (-6.39, 7.48) | 0.932 |
| Continuous variable  (10 μg/m^3^ PM_2.5_ increment) | Unadjusted | 2.03 (-1.21, 5.36) | 0.222 |
|  | Standard adjustment^*^ | 1.50 (-1.76, 4.87) | 0.371 |

^*^ The standard model included the covariates of annual temperature, urban/rural residency, sex, educational level, marriage status, smoking, and drinking.

Table S2. Estimated associations of mental health with PM_2.5_ concentration and age using mixed-effects models.

| Model settings | | Excess risk (%) with 95% confidence intervals | | |
| --- | --- | --- | --- | --- |
|  |  | PM_2.5_ concentration (per 10 μg/m^3^) | Age (per year) | Interaction |
| Unadjusted model | PM_2.5_ concentration only | 1.28 (0.11, 2.47) |  |  |
|  | Age only |  | 0.53 (0.42, 0.63) |  |
|  | PM_2.5_ concentration and age | 3.50 (1.89, 5.12) | 0.91 (0.59, 1.22) | -0.06 (-0.11, -0.01) |
| Adjusted model | Standard^*^ | 3.63 (2.00, 5.27) | 0.76 (0.45, 1.07) | -0.06 (-0.11, -0.01) |
|  | Fully adjusted^#^ | 3.07 (1.44, 4.73) | 0.75 (0.45, 1.06) | -0.07 (-0.12, -0.02) |

^*^ The standard model included the covariates of annual temperature, urban/rural residency, sex, educational level, marriage status, smoking, and drinking.

^#^ The fully adjusted model further included the covariates of cooking energy type, building type, residential rent payment, presence of an in-house bath facility, presence of an in-house telephone, and indoor temperature maintenance.

Table S3. Comparison of the random- and fixed-effects models.

| Variable | Reference (for categorical variables) or unit (for continuous variables) | Excess risk (%)^#^ | |
| --- | --- | --- | --- |
|  |  | Random-effects model | Fixed-effects model |
| PM_2.5_ | 10 μg/m^3^ | 3.63 (2.00,5.27) | 4.21 (1.48,7.02) |
| Age | 1 year | 0.76 (0.45,1.07) | 0.45 (-0.34,1.24) |
| PM_2.5_*age | (10 μg/m^3^) * (1 year) | -0.06 (-0.11,-0.01) | -0.23 (-0.35,-0.12) |
| Urban | Rural | -10.52 (-13.81,-7.09) |  |
| Female | Male | -19.57 (-21.58,-17.50) |  |
| Elementary & middle | Below elementary | -9.43 (-11.46,-7.35) |  |
| Above middle |  | -19.59 (-22.34,-16.75) |  |
| Married | Unmarried | -11.73 (-13.74,-9.68) | -9.87 (-13.01,-6.61) |
| Smoking | Non-smoking | 4.53 (1.92,7.20) | -0.00 (-10.61,11.85) |
| Never drinking | Frequently drinking | 4.97 (2.74,7.26) | 0.11 (-2.95,3.27) |
| Rarely drinking |  | 3.03 (0.04,6.11) | 0.75 (-2.80,4.42) |

\# Standard adjustment (Table S1) was applied to both the random- and fixed-effects models.

Table S4. Longitudinal cross-sectional estimates of the effect of age.

| Model | Excess risk (%) for per year increment of age | | | |
| --- | --- | --- | --- | --- |
|  | Longitudinal estimate^*^ | Cross-sectional estimates^*^ | | |
|  |  | CHARLS 2011 | CHARLS 2013 | CHARLS 2015 |
| Unadjusted model^#^ | 0.53 (0.42, 0.63) | 0.66 0.57 0.75 | 0.60 0.51 0.69 | 0.56 0.47 0.64 |
| Standard adjustment model^#^ | 0.76 (0.45, 1.07) | 0.76 (0.49, 1.02) | 0.71 (0.44, 0.97) | 0.65 (0.39, 0.91) |

^*^ The longitudinal model and the corresponding cross-sectional model were identical, with the exception of the following: (1) the cross-sectional model lacked the subject-specific random slope; and (2) the cross-sectional model estimated the effect of age by survey wave.

^#^ The details of the model adjustment are provided in the footnotes to Table S1.

Table S5 The within-city variance in PM_2.5_.

|  | Percentage of within-city variance in total variance of PM_2.5_ (2011 or 2015) or PM_2.5_ reduction (2011-2015)^*^ | | |
| --- | --- | --- | --- |
|  | 2011 | 2015 | 2011-2015 |
| All cities^#^ | 35.84% | 38.42% | 17.89% |
| Studied cities | 17.60% | 20.16% | 11.75% |

* The percentage is calculated from the gridded maps of annual PM_2.5_ concentrations, with a spatial resolution of 0.1° × 0.1°. The distribution for total, between-city or within-city variation of PM_2.5_ is displayed in Figure S8.

\# All 345 cities in the mainland of China.

**Supplementary codes**

###R code of the main regression models in the following paper###

##Authors: Tao Xue, Tianjia Guan, Yixuan Zheng, Guannan Geng, Qiang Zhang, Tong Zhu##

#The code is provided as a demo (using the standard adjustment model). Please don't run it directly.#

#For more information, please contact Dr. Tao Xue#

#Load the package for statistical analysis#

library(lme4)

library(splines)

library(plm)

library(ipw)

#Load the main database, which is publicly available from the CHARLS team (http://opendata.pku.edu.cn/).#

#However, we don't have the permission to re-distribute the CHARLS data, according to the user's guidelines.#

load("dta.RData")

#Longitudinal model: random effect#

m<-lmer(log(score+1) ~ PM25 + ns(TMP,3) + ages + I(PM25*ages) + urban_nbs + sex + edu + married + smoke + drink + city2 + (1|site) + (1|ID2), data=dta)

#Longitudinal model: fixed effect#

m<-plm(log(score+1) ~ PM25 + ns(TMP,3) + ages + I(PM25*ages) + urban_nbs + sex + edu + married + smoke + drink, index=c("ID2","vis"), data=dta, model="within")

#Difference-in-difference model#

#Load database for difference-in-difference analysis, which is derived from the CHARLS 2011 and 2015

load("dd dta.RData")

ws<-ipwpoint(exposure=PM25,family = "gaussian",

numerator = ~ 1,denominator=~urban_nbs + sex + edu + age + TMP, data=dd)

dd$ws=ws$ipw.weights

#Difference-in-difference model#

m<-lmer(y ~ PM25 + ns(age,3) + urban_nbs + sex + edu + ns(TMP,3) + married + smoke + drink + (1|site) + (1|HID),

data=dd, weights=ws)

#Results interpretation for all models: excess risk (%) with 95% CIs for PM2.5 (per 10 ug/m3)

ER = (exp(summary(m)$coef["PM25","Estimate"] * 10) - 1) * 100

ER.CI.lo = (exp((summary(m)$coef["PM25", "Estimate"] – 1.96*summary(m)$coef["PM25", "Std. Error"]) * 10) - 1) * 100

ER.CI.up = (exp((summary(m)$coef["PM25", "Estimate"] + 1.96*summary(m)$coef["PM25", "Std. Error"]) * 10) - 1) * 100
